# Associations of Cardiovascular-Kidney-Metabolic syndrome with premature mortality and life expectancies in US adults: a cohort study

**DOI:** 10.1101/2024.12.19.24319121

**Authors:** Lubi Lei, Jingkuo Li, Wei Wang, Yanwu Yu, Boxuan Pu, Yue Peng, Lihua Zhang, Yuanlin Guo

## Abstract

**Background:** The American Heart Association recently updated the Cardiovascular-Kidney-Metabolic (CKM) Health advisory, proposing a new framework for defining, staging, and predicting CKM risk. However, the prevalence and adverse effects of CKM stages remain insufficiently characterized.

**Methods:** We analyzed data from the National Health and Nutrition Examination Survey (NHANES) (1999–2018), including 18,350 US adults aged 20– 79 years. CKM syndrome encompasses subclinical or clinical cardiovascular disease (CVD), chronic kidney disease (CKD), and metabolic risk factors. Participants were categorized into four CKM stages based on clinical severity. We assessed associations of CKM stages with mortality risk and life expectancy.

**Results:** Only 12.9% of participants were classified as CKM stage 0. The prevalence of CKM stages 1, 2, 3, and 4 was 23.1%, 53.6%, 3.6%, and 6.7%, respectively. Compared to CKM stage 0, individuals in stage 4 had a markedly higher risk of all-cause mortality (HR: 4.30, 95% CI: 2.95– 6.26) and lost 15.5 (12.5–19.8) years of life at age 50. Sex and racial/ethnic disparities were also observed.

**Conclusions:** Higher CKM stages were strongly associated with increased mortality and reduced life expectancy. Our findings underscore the urgent need for enhanced CKM health management, social support, and policy interventions.

**Key Messages:** *What is already known on this topic:*

- Metabolic disorders, chronic kidney disease, and cardiovascular diseases are significantly associated with premature mortality. By taking into account the overlaps between cardiovascular-kidney-metabolic (CKM) syndrome, it is possible to improve risk stratification and employ an integrated approach to multimorbidity management.

*What this study adds?:*

- Only 12.6% of individuals had no CKM conditions, whereas approximately 50% of individuals had CKM stage 2, encompassing metabolic disorders, chronic kidney disease, or a combination of both.
- The estimated life expectancy of individuals with CKM stage 4 was 15.5 years lower than that of individuals with CKM stage 0 at the age of 50.

*How this study might affect research, practice or policy?:*

- Our results indicate that the CKM condition is prevalent among US adults. We advocate the implementation of the new staging system for the prevention and treatment of CKM syndrome.
- Life expectancy is reduced in adults who have a combination of metabolic risk factors, chronic kidney disease, or cardiovascular disease. We highlight the importance of incorporating a people-centered approach that includes the development of social support and policy, the integration of CKM health multidisciplinary management, and the enhancement of obesity management.

## Introduction

Poor cardiovascular-kidney-metabolic (CKM) health is a significant issue of premature mortality and excess morbidity in the US, as well as substantial social and economic burdens.^1, 2^ CKM syndrome is a composite concept that refers to the clinical manifestation of interactions between metabolic risk factors, chronic kidney disease (CKD) and cardiovascular disease (CVD).^3^ The syndrome is prevalent and accounts for approximately 26.3% of the general population, which has substantially increased over the past two decades.^4^ Physical and mental impairments are typically present with CKM conditions.^2^ Given the clinical and public health priorities, there is an increasing necessity to further recognize the prevalence of varying degrees of CKM severity and the corresponding health outcomes.

The American Heart Association recently released an updated advisory for the definition of CKM conditions, as well as its staging and risk prediction approach.^3, 5^ Prior studies suggested that the mortality rate attributed to either component of CKM syndrome has increased over time.^6–8^ Nevertheless, to the best of our knowledge, no studies have conducted a comprehensive evaluation of the new staging system in relation to health outcomes. Efforts should be paid to promote public awareness of the current recommendations for improving CKM health in the general population.

In this study, we aimed to describe the prevalence of CKM stages and various combinations of conditions in each stage and to assess the associations between CKM stages and the risk of mortality and life expectancy among US adults by utilizing data from the National Health and Nutrition Examination Survey (NHANES).

## Materials and Methods

### Patient and Public Involvement statement

Patients and/or the public were not involved in the design, or conduct, or reporting, or dissemination plans of this research.

### Study population

We used data from ten NHANES cycles from 1999 to 2018 (downloaded at http://www.cdc.gov/nchs/nhanes.htm).^9^ The NHANES is a national survey with a complex, 4-stage probability sampling design to select a representative sample of the civilian, noninstitutionalized US population. The NHANES was approved by the National Center for Health Statistics Research Ethics Review Board and obtained written informed consent from all participants or their parents or guardians.^10^ In the mobile examination center (MEC) examination, a total of 22149 nonpregnant participants aged 20 to 79 years participated in the fasting test. We excluded 3176 participants with insufficient information to ascertain CKM syndrome, 27 with missing death data, and 596 participants with missing covariates. This resulted in a final sample of 18350 participants with available data (**eFigure 1**).

### Assessments of CKM

Information was collected via a standardized questionnaire and physical examinations through face-to-face interviews. Blood and random urine samples were analysed in the central laboratory. We calculated the estimated glomerular filtration rate (eGFR) through the 2021 race- and ethnicity-free Chronic Kidney Disease Epidemiology Collaboration creatinine equation.^11^

CKM syndrome was defined as the presence or co-occurrence of subclinical or clinical CVD, CKD, metabolic risk factors or excess/dysfunctional adiposity. Any history of chronic heart failure, coronary heart disease, heart attack, or stroke was considered clinical cardiovascular disease (CVD). Subclinical CVD was defined as having a 10-year CVD risk of >= 20% or very high-risk CKD. We calculated the predicted 10-year CVD risk using the PREVENT score, which includes age, sex, tobacco use, blood pressure, cholesterol, diabetes, kidney function, antihypertensive agents use, and statin use in the model (algorithm was detailed in **eTable 1**).^12^ We classified risks of CKD based on the Kidney Disease: Improving Global Outcomes (KDIGO) classification,^13^ which includes various thresholds for eGFR (<30, 30-44, 45-59,≥60 ml/min/1.73m^2^) and urinary albumin creatinine ratio (UACR, <30, 30-299, ≥300 mg/g). Those with moderate or high risk were classified as having CKD and very high-risk CKD was classified as having subclinical CVD.^14^ Metabolic risk factors include diabetes, hypertension, dyslipidemia and metabolic syndrome (MetS).

Excess/dysfunctional adiposity encompasses prediabetes, overweight/obesity measured by BMI, and abdominal obesity measured by waist circumference. We elected to categorize participants according to the four CKM stages. Stage 0 refers to all normal conditions. Stage 1 is characterized by only excess/dysfunctional adiposity. Stage 2 is characterized by at least one of metabolic risk factors and/or CKD. Stage 3 is characterized by subclinical CVD with excess/dysfunctional adiposity, metabolic risk factors or CKD. Stage 4 is characterized by CVD with excess/dysfunctional adiposity, metabolic risk factors, CKD, or very high CKD risk.^3^ The detailed stage criteria are provided in **eTable 2**.

### Assessments of other covariates

Other covariates in this study included age, sex (male, female), and race/ethnicity (non-Hispanic White, non-Hispanic Black, Mexican, other Hispanic and other races [including Asian or multiracial]). We refrained from combining Mexican individuals and other Hispanic individuals in this study because the NHANES oversampled Mexican individuals before 2007 rather than all Hispanic individuals.^15^ We defined employed, high-income level, full food security, high education attainment, private insurance, owning house, and partnered as the positive social determinants of health (SDOH) domain,^16^ which was detailed in **eTable 3**. The SDOH score was the number of positive SDOH domains, which ranged from 0 to 8. CKM risk factors included hemoglobin a1c (HbA1c), total cholesterol (TC), UACR, eGFR and self-reported diagnosis history of cancer, liver disease and lung disease.

### Ascertainment of death

The clinical outcomes in this study included all-cause death and CV death. Mortality data was ascertained via linkage and probabilistic record matching with the National Death Index (NDI) through 31 December 2019.^17^ CV deaths were defined as deaths due to major CVD or cerebrovascular diseases (International Classification of Diseases, Tenth Edition [ICD-10] codes: I00-I09, I11, I13, I20-I51 and I60-I69). The time to event was counted from the interview time to the date of death or 31 December 2019, whichever came first.

To determine life expectancy, we obtained the population all-cause mortality rate from 50 to 100 years by single-year age in 2019 from the National Center for Health Statistics (NCHS). We also derived the population CV mortality rates from 50 to 84 years by single-year age in 2019 from the CDC (Centers for Disease Control and Prevention) WONDER (Wide-Ranging Online Data for Epidemiologic Research) database of the US population.^18^ Because the database did not contain any CV death rates for individuals older than or equal to 85 years, we estimated single-year CV death rates from 85 to 100 years by extrapolation using the Poisson regression model with log-linear and quadratic terms for the sequence from 50 to 100 years (**eFigure 2**).

### Statistical analysis

According to analytic guidelines published by the NCHS, stratum and primary sampling units were considered for the complex, multistage, probability sampling design.^19^ All analyses employed examination sample weights, encompassing those who participated in an MEC examination. We described continuous variables by means with weighted 95% confidence intervals (CIs) and categorical variables by frequencies with weighted percentages. We calculated the age-adjusted prevalence of CKM stages by the direct method to the 2000 US Census population using three age groups (20-39, 40-59, and ≥ 60 years).

After we assessed the proportional hazards assumption, we examined associations between CKM stages and the risk of death using survey Cox proportional hazards regression with examination weights as weights, primary sampling units as clusters and secondary sampling units as stratifications. Since the survey Cox proportional hazards regression model was unable to perform competing risk analyses, non-CV death was censored in models with CV death. We conducted the Fine-Gray subdistribution hazard model to account for the competing risk of non-CV death as a sensitivity analysis. Models were adjusted for age, sex, race/ethnicity, and SDOH score. Moreover, to examine whether age, sex, race/ethnicity and SDOH differed, we conducted stratified analyses by age (20-34, 35-49, 50-64, and ≥ 65 years), sex, race/ethnicity groups (White, Black, and Mexican) or SDOH score (=8, <8). We also incorporated multiplicative interaction terms into the models.

We calculated the life expectancy of participants with different CKM phases using the life table method.^20^ We developed a life table that includes one-year age bands for the ages of 50 to 100. The cumulative survival from age 50 onwards was estimated for participants with different CKM stages, by weighting the adjusted hazard ratios (HRs) for all-cause death from the NHANES, the age-specific population all-cause death rate in 2019, and the age-specific weighted prevalence of each CKM stage from the NHANES. The years of life loss in stage 1-4 were estimated as the difference in life expectancy at age 50 between stage 0 and the other stages. The methods used for estimating life expectancy are detailed in **eMethods 1**. By applying Arriaga’s decomposition method,^21^ we estimated the CVD-cause contributions to the life expectancy difference between participants with stage 4 and stage 0 (detailed in **eMethods 2**). Given that life expectancy varied by sex and race/ethnicity, we performed the corresponding stratified analyses. We did not perform the analyses in SDOH subgroups due to the limited information available on SDOH-specific population death rates in the US. In both the Cox models and the life expectancy calculation, we performed a sensitivity analysis that further adjusted for CKM risk factors (i.e., HbA1c, TC, UACR, eGFR and self-reported diagnosis history of cancer, liver disease and lung disease).

All analyses were conducted using SAS 9.4 software (SAS Institute, Cary, NC) and R 4.4.2 (R Foundation for Statistical Computing, Vienna, Austria). We calculated the confidence interval (CI) for life expectancy by bootstrapping method with 1000 runs. A two-sided P value <0.05 was considered significant.

## Results

### Baseline characteristics

In our study, 18350 participants aged 20-79 years were included, representing 79 945 838 noninstitutionalized US adults. The average age was 46.0 years (95% CI: 45.6, 46.5), 51.0% of the participants were female (n=9398), and 68.9% of the participants were non-Hispanic White (n=7920). Compared with those in stage 0, participants in stage 4 were older, more likely to be male, and had lower SDOH scores (**Table 1 and eTable 4**). Compared with the study sample, participants who had incomplete information were more likely to be non-Hispanic Black and to have lower SDOH scores (**eTable 5**). There were 1873 (age-adjusted 12.9%) participants in CKM stage 0, 3870 (age-adjusted 23.1%) in stage 1, 9960 (age-adjusted 53.6%) in stage 2, 1103 (age-adjusted 3.6%) in stage 3, and 1544 (age-adjusted 6.7%) in stage 4. Females had a higher prevalence of CKM stage 0-1 than males (**Figure 1**). The age-adjusted prevalences by SDOH, race/ethnicity and survey cycle subgroups were shown in **eFigure 3-5**.

**Table 1.**
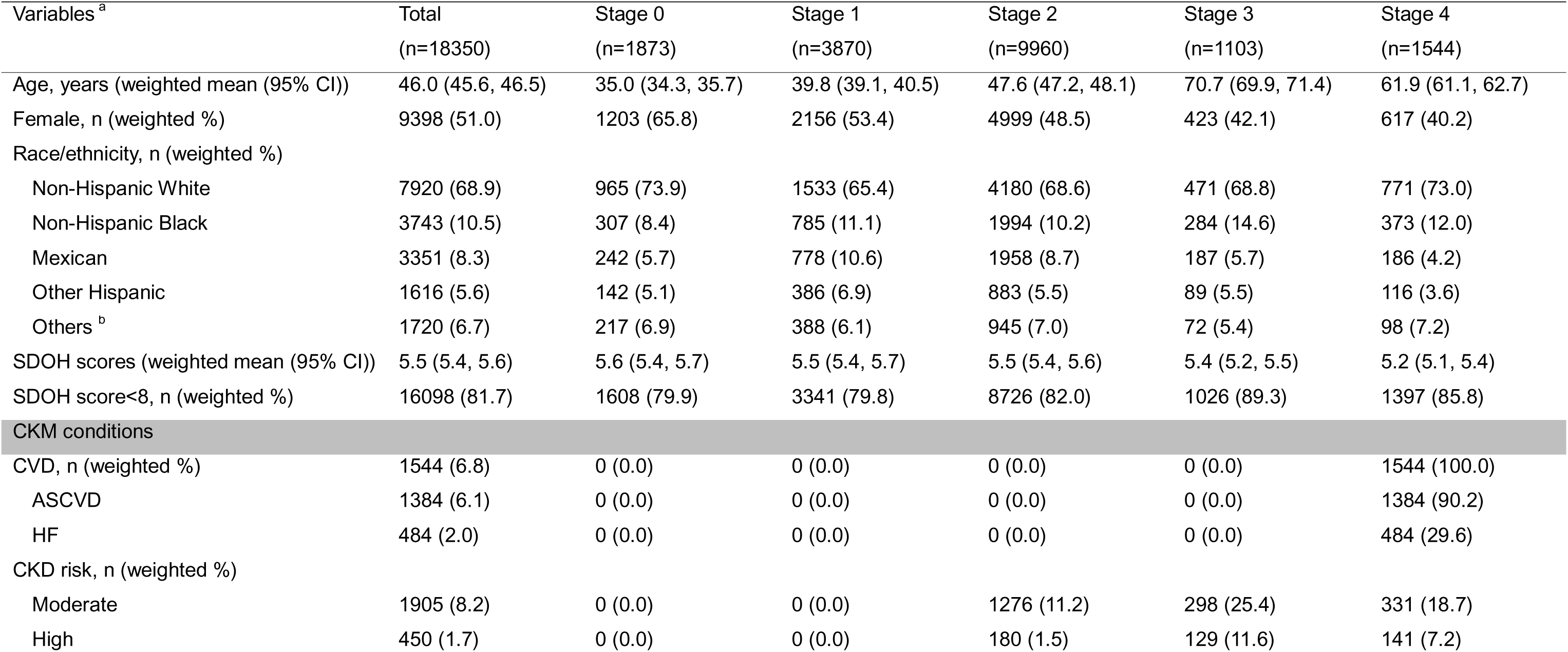
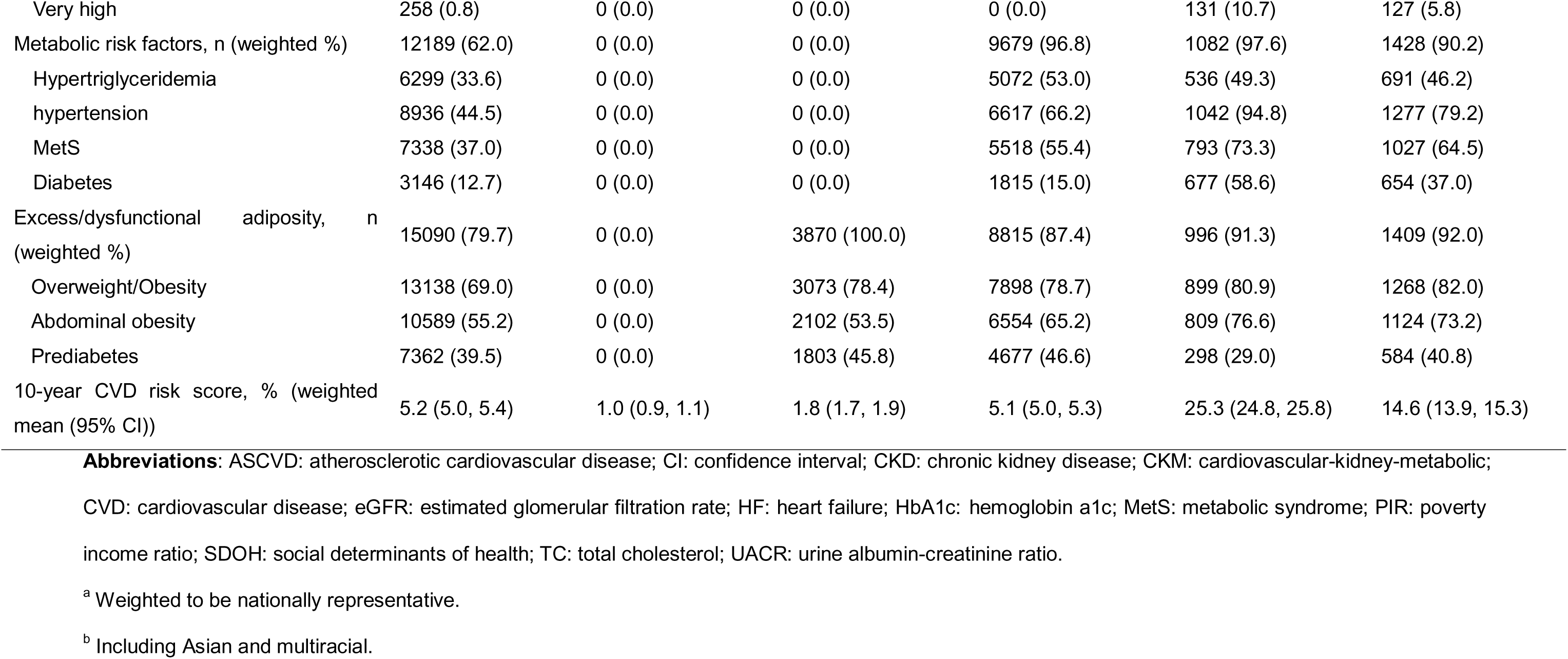
Sample size and key characteristics by CKM stages.

**Figure 1.**
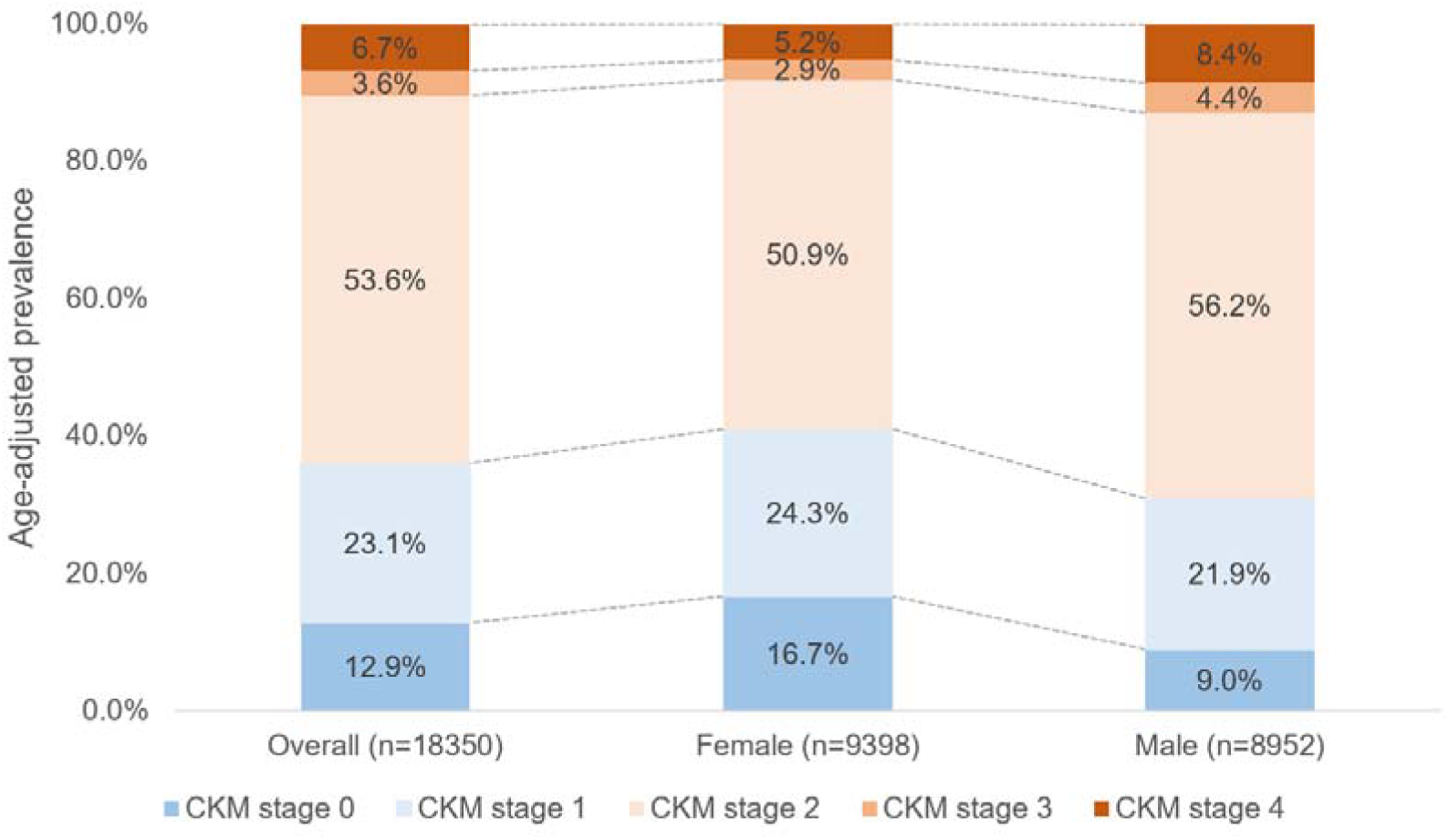
Age adjusted prevalence of CKM stages in overall individuals and by sex subgroups **Abbreviations:** CKM: cardiovascular-kidney-metabolic.

### Prevalence and overlaps of CKM conditions in each stage

Excess/dysfunctional adiposity was the most prevalent CKM condition (weighted 79.7%), followed by metabolic risk factors (weighted 62.0%), CKD (weighted 9.9%), CVD (weighted 6.8%) and subclinical CVD (weighted 5.3%) (**Table 1**). In stage 1, the most common CKM condition was obesity only (weighted 54.2%). In stage 2, “obesity + metabolic risk factors” (weighted 36.2%) and “obesity + prediabetes + metabolic risk factors” (weighted 35.3%) were disproportionally high. In stage 3, “obesity + metabolic risk factors + high-risk CVD” (weighted 33.2%) and “obesity + metabolic risk factors + high-risk CVD + moderate-to-high CKD” (weighted 20.4%) were the top two combinations. In stage 4, “obesity + metabolic risk factors + CVD” (weighted 23.2%) and “obesity + prediabetes + metabolic risk factors + CVD” (weighted 20.4%) were the top two combinations (**Table 2**). The prevalence of each stage varied between groups when stratified by sex (**Table 2**), survey cycles (**eTable 6**), and race/ethnicity (**eTable 7**). Nevertheless, the most prevalent combination of conditions in each stage remained consistent across subgroups.

**Table 2.**
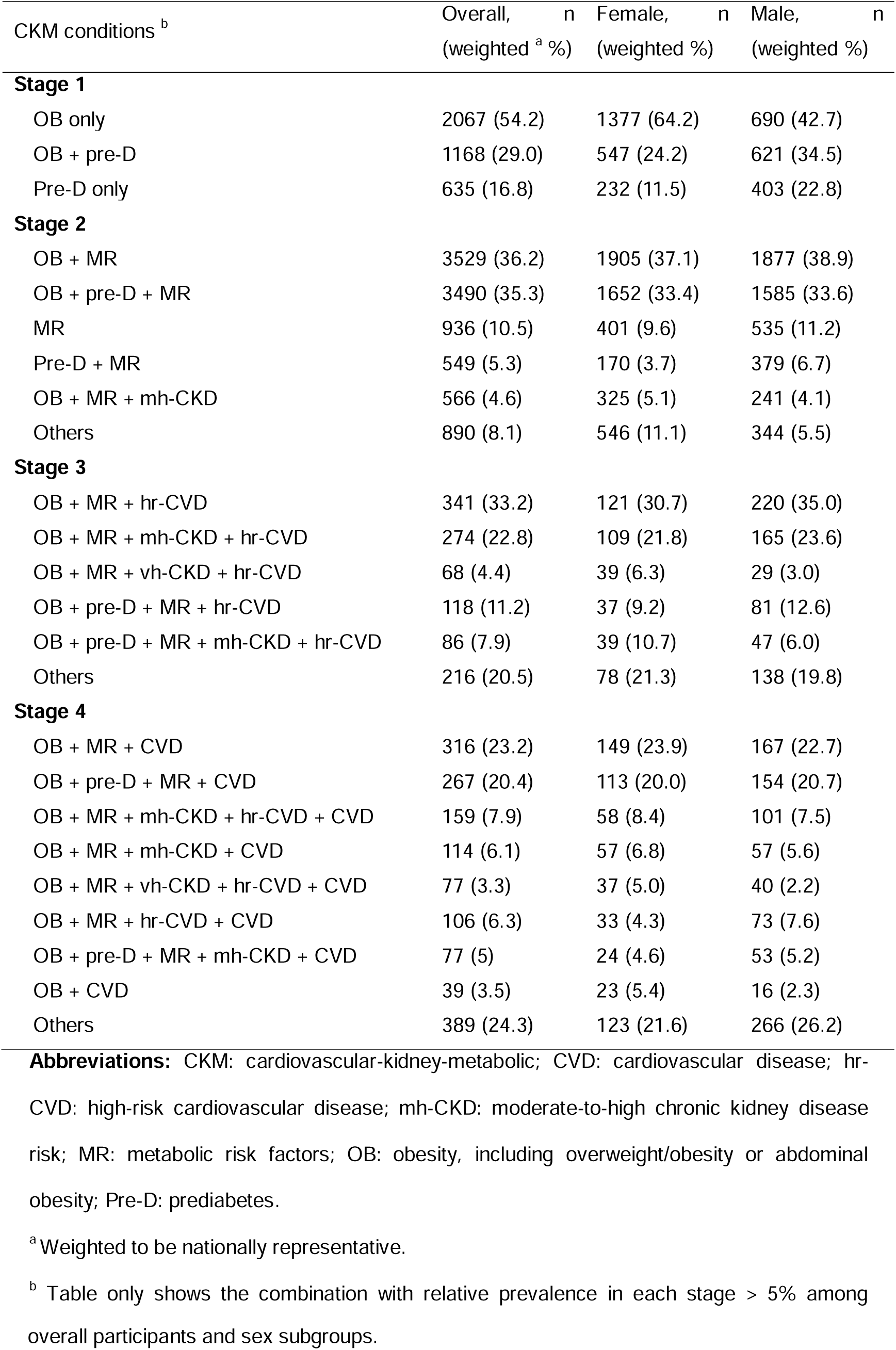
Relative proportions of different combinations of CKM conditions stratified by CKM stage 1-4.

| CKM conditions <sup>b</sup> | Overall, n<br>(weighted <sup>a</sup> %) | Female, n<br>(weighted %) | Male, n<br>(weighted %) |
| --- | --- | --- | --- |
| **Stage 1** |  |  |  |
| OB only | 2067 (54.2) | 1377 (64.2) | 690 (42.7) |
| OB + pre-D | 1168 (29.0) | 547 (24.2) | 621 (34.5) |
| Pre-D only | 635 (16.8) | 232 (11.5) | 403 (22.8) |
| **Stage 2** |  |  |  |
| OB + MR | 3529 (36.2) | 1905 (37.1) | 1877 (38.9) |
| OB + pre-D + MR | 3490 (35.3) | 1652 (33.4) | 1585 (33.6) |
| MR | 936 (10.5) | 401 (9.6) | 535 (11.2) |
| Pre-D + MR | 549 (5.3) | 170 (3.7) | 379 (6.7) |
| OB + MR + mh-CKD | 566 (4.6) | 325 (5.1) | 241 (4.1) |
| Others | 890 (8.1) | 546 (11.1) | 344 (5.5) |
| **Stage 3** |  |  |  |
| OB + MR + hr-CVD | 341 (33.2) | 121 (30.7) | 220 (35.0) |
| OB + MR + mh-CKD + hr-CVD | 274 (22.8) | 109 (21.8) | 165 (23.6) |
| OB + MR + vh-CKD + hr-CVD | 68 (4.4) | 39 (6.3) | 29 (3.0) |
| OB + pre-D + MR + hr-CVD | 118 (11.2) | 37 (9.2) | 81 (12.6) |
| OB + pre-D + MR + mh-CKD + hr-CVD | 86 (7.9) | 39 (10.7) | 47 (6.0) |
| Others | 216 (20.5) | 78 (21.3) | 138 (19.8) |
| **Stage 4** |  |  |  |
| OB + MR + CVD | 316 (23.2) | 149 (23.9) | 167 (22.7) |
| OB + pre-D + MR + CVD | 267 (20.4) | 113 (20.0) | 154 (20.7) |
| OB + MR + mh-CKD + hr-CVD + CVD | 159 (7.9) | 58 (8.4) | 101 (7.5) |
| OB + MR + mh-CKD + CVD | 114 (6.1) | 57 (6.8) | 57 (5.6) |
| OB + MR + vh-CKD + hr-CVD + CVD | 77 (3.3) | 37 (5.0) | 40 (2.2) |
| OB + MR + hr-CVD + CVD | 106 (6.3) | 33 (4.3) | 73 (7.6) |
| OB + pre-D + MR + mh-CKD + CVD | 77 (5) | 24 (4.6) | 53 (5.2) |
| OB + CVD | 39 (3.5) | 23 (5.4) | 16 (2.3) |
| Others | 389 (24.3) | 123 (21.6) | 266 (26.2) |

**Abbreviations:** CKM: cardiovascular-kidney-metabolic; CVD: cardiovascular disease; hr-CVD: high-risk cardiovascular disease; mh-CKD: moderate-to-high chronic kidney disease risk; MR: metabolic risk factors; OB: obesity, including overweight/obesity or abdominal obesity; Pre-D: prediabetes.
<sup>a</sup>Weighted to be nationally representative.
<sup>b</sup> Table only shows the combination with relative prevalence in each stage > 5% among
overall participants and sex subgroups.

### Association between CKM stages and risk of death

During the median follow-up of 9.6 (interquartile range: 5.3, 14.1) years, 2054 (11.2%) participants died, and 604 (3.3%) died of CV causes. Compared with participants in stage 0, participants in stage 2 had an increased risk of all-cause death (HR: 1.68, 95% CI: 1.20, 2.35) and CV death (HR: 3.46, 95% CI: 1.17, 10.20). Similarly, participants in stage 3 and stage 4 had increased risks of all-cause death (stage 3: HR: 3.65, 95% CI: 2.49, 5.35; stage 4: HR: 4.30, 95% CI: 2.95, 6.26) and CV death (stage 3: HR: 9.85, 95% CI: 3.11, 31.20; stage 4: HR: 13.30, 95% CI: 4.45, 39.70). No significant association was observed between stage 1 and all-cause death (HR: 1.14, 95% CI: 0.72, 1.80) or CV death (HR: 1.75, 95% CI: 0.56, 5.48) (**Table 3**). Similar associations between CKM stages and the risk of mortality were observed after further stratification by sex (**Table 3**), age (**eTable 8**), SDOH score (**eTable 9**), and race/ethnicity subgroups (**eTable 10**). The associations of higher CKM stages with the risk of mortality were stronger in non-Hispanic White individuals, older individuals and those who did not meet all SDOH-positive domains than in their counterparts. We did not observe significant interactions with CKM stages in any subgroup (P for interaction>0.05). The results were comparable to those of the main analyses following additional adjustments for CKM risk factors (**eTable 11**). The results were consistent with the main analyses after employing the competing risk model to account for non-CV death. The point estimates were attenuated, particularly among the participants aged 60-79 years and the Black participants (**eTable 12**).

**Table 3.**
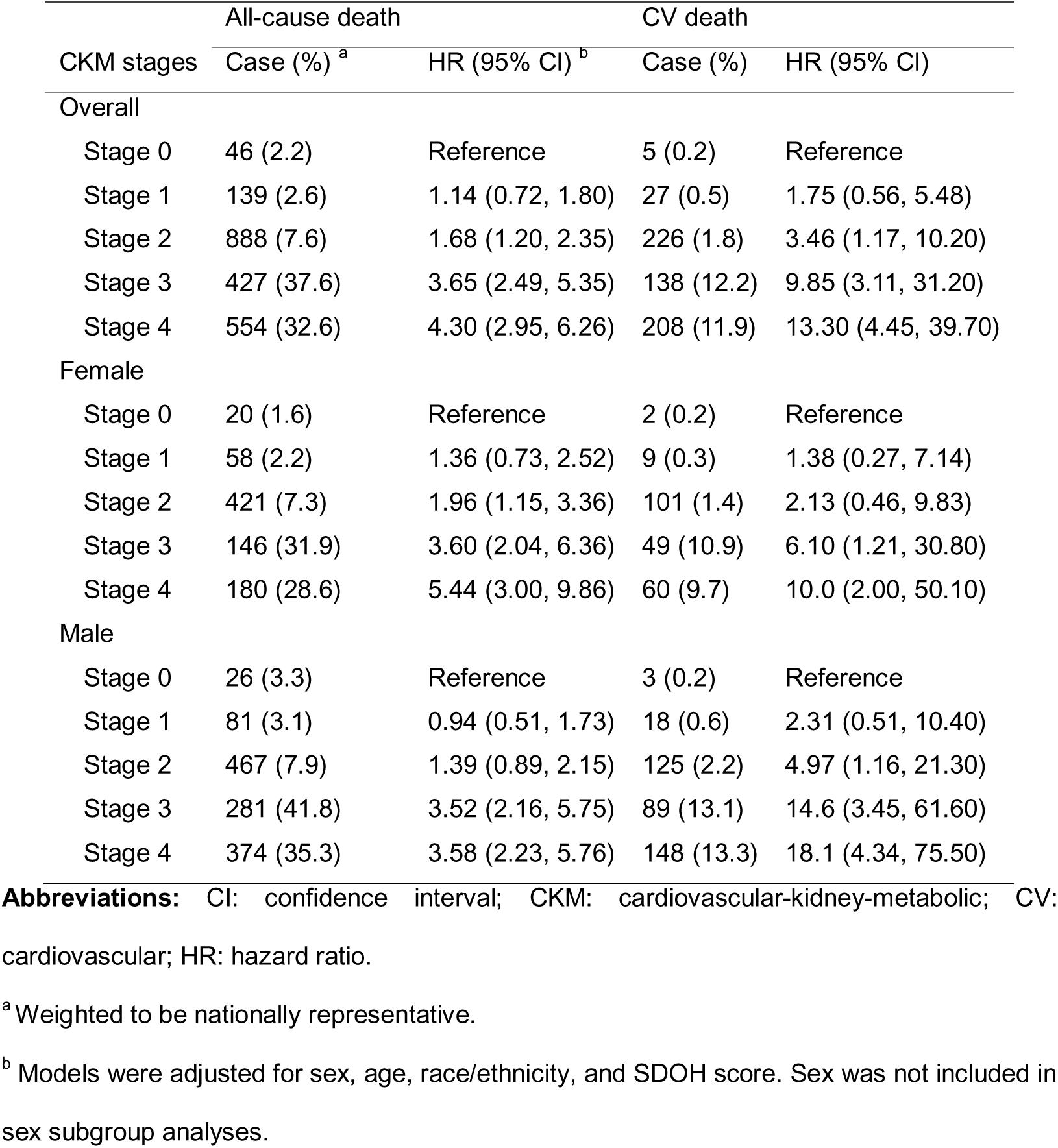
Associations of CKM stages with all-cause death and cardiovascular death in overall individuals and by sex subgroups.

| CKM stages | All-cause death |  | CV death |  |
| --- | --- | --- | --- | --- |
|  | Case (%) <sup>a</sup> | HR (95% CI) <sup>b</sup> | Case (%) | HR (95% CI) |
| Overall |  |  |  |  |
| Stage 0 | 46 (2.2) | Reference | 5 (0.2) | Reference |
| Stage 1 | 139 (2.6) | 1.14 (0.72, 1.80) | 27 (0.5) | 1.75 (0.56, 5.48) |
| Stage 2 | 888 (7.6) | 1.68 (1.20, 2.35) | 226 (1.8) | 3.46 (1.17, 10.20) |
| Stage 3 | 427 (37.6) | 3.65 (2.49, 5.35) | 138 (12.2) | 9.85 (3.11, 31.20) |
| Stage 4 | 554 (32.6) | 4.30 (2.95, 6.26) | 208 (11.9) | 13.30 (4.45, 39.70) |
| Female |  |  |  |  |
| Stage 0 | 20 (1.6) | Reference | 2 (0.2) | Reference |
| Stage 1 | 58 (2.2) | 1.36 (0.73, 2.52) | 9 (0.3) | 1.38 (0.27, 7.14) |
| Stage 2 | 421 (7.3) | 1.96 (1.15, 3.36) | 101 (1.4) | 2.13 (0.46, 9.83) |
| Stage 3 | 146 (31.9) | 3.60 (2.04, 6.36) | 49 (10.9) | 6.10 (1.21, 30.80) |
| Stage 4 | 180 (28.6) | 5.44 (3.00, 9.86) | 60 (9.7) | 10.0 (2.00, 50.10) |
| Male |  |  |  |  |
| Stage 0 | 26 (3.3) | Reference | 3 (0.2) | Reference |
| Stage 1 | 81 (3.1) | 0.94 (0.51, 1.73) | 18 (0.6) | 2.31 (0.51, 10.40) |
| Stage 2 | 467 (7.9) | 1.39 (0.89, 2.15) | 125 (2.2) | 4.97 (1.16, 21.30) |
| Stage 3 | 281 (41.8) | 3.52 (2.16, 5.75) | 89 (13.1) | 14.6 (3.45, 61.60) |
| Stage 4 | 374 (35.3) | 3.58 (2.23, 5.76) | 148 (13.3) | 18.1 (4.34, 75.50) |

**Abbreviations:** CI: confidence interval; CKM: cardiovascular-kidney-metabolic; CV:
cardiovascular; HR: hazard ratio.
<sup>a</sup> Weighted to be nationally representative.
<sup>b</sup> Models were adjusted for sex, age, race/ethnicity, and SDOH score. Sex was not included in sex subgroup analyses.

### CKM stages and life expectancy

At the age of 50, the estimated life expectancies for participants in stages 0, 1, 2, 3, and 4 were 41.4 years (95% CI: 38.6, 45.5), 40.0 years (95% CI: 38.3, 42.2), 35.8 years (95% CI: 35.1, 36.5), 27.6 years (95% CI: 26.6, 28.6), and 25.9 years (95% CI: 25.2, 26.7), respectively. We observed a dose-response relationship between CKM stages and years of life loss. Participants in stage 4 lost more than 15.5 years (95% CI: 12.5, 19.8) at age 50 than those in stage 0. There was no significant difference in life expectancy between stage 0 and stage 1 (years: 1.6, 95% CI: −1.8, 5.5) (**Figure 2**). In stage 4, females (16.9 years) and Mexicans (17.5 years) experienced greater life loss than their counterparts (male: 14.1 years; White individuals: 15.2 years; Black individuals: 15.0 years) (**Figure 2**). **eFigure 6** shows the aggregate life expectancy of the sex and race/ethnicity subgroups in stages 0 to 4. The results were comparable to those of the main analyses following additional adjustments for CKM risk factors (**eFigure 7**).

**Figure 2.**
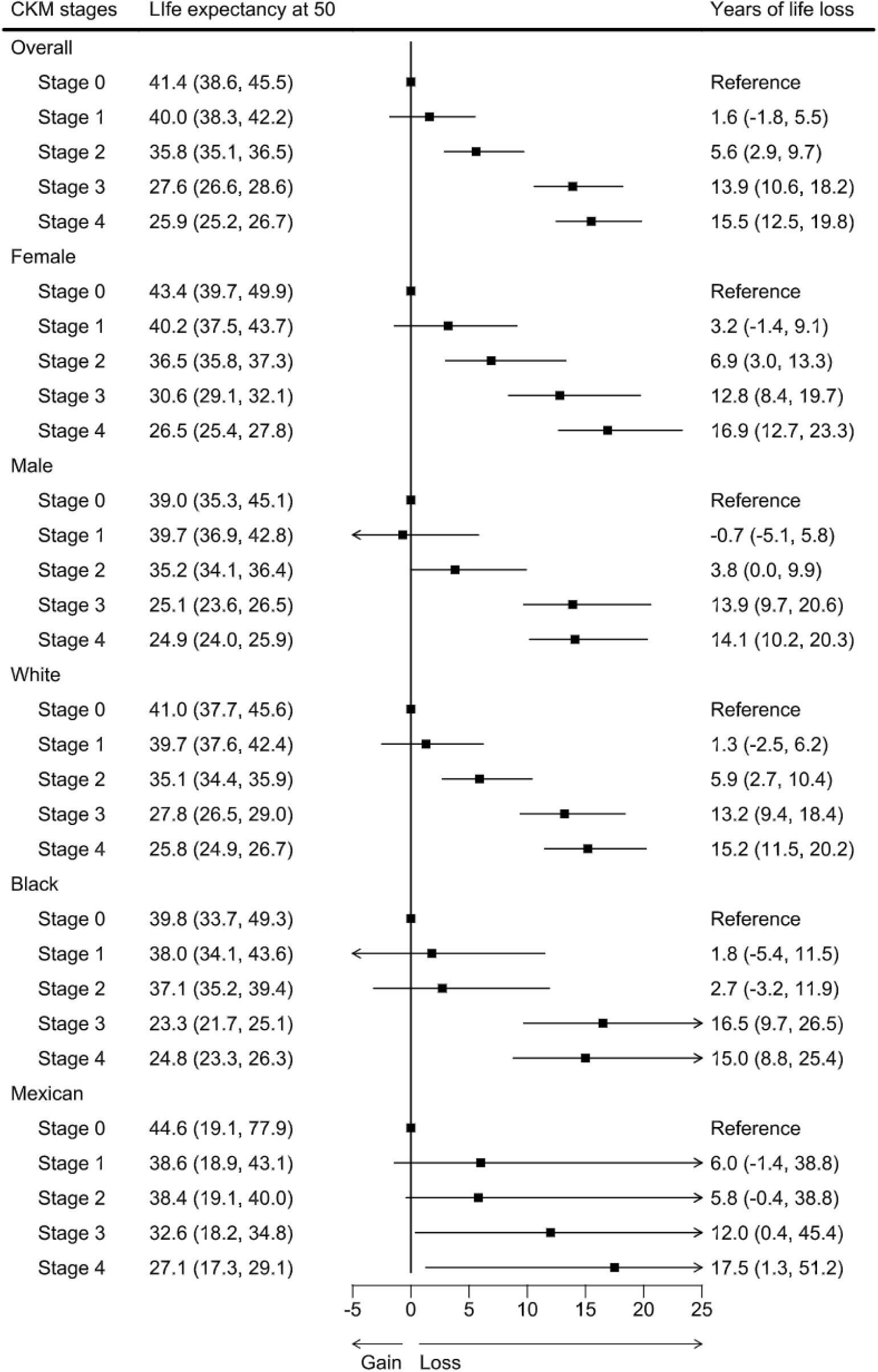
Life expectancy and years of life loss at age 50 according to CKM stages in overall individuals and by sex and race/ethnicity subgroups **Abbreviations:** CKM: cardiovascular-kidney-metabolic.

An average of 39.6% of the lost life expectancy at age 50 years from stage 4 was attributable to CV death (**eFigure 8**). The percentage was higher in male (45.3%) and Black individuals (48.1%) than in their counterparts (female: 32.3%; White individuals: 39.5%; Mexican individuals: 26.6%) (**eFigure 8-9**). The detailed life expectancies across ages 50 to 100 in each stage for overall individuals and by sex subgroups were presented in **eTable 13** and race/ethnicity subgroups are presented in **eTable 14**.

## Discussion

By leveraging a large, nationally representative sample of US adults, we found that over half of individuals had comorbidities of obesity and metabolic abnormalities. Individuals with a higher stage of CKM syndrome had an increased risk of all-cause death and CV death. The overall decrease in life expectancy in CKM stage 4 was approximately 40% which was attributable to CV death. Disparities existed in sex and race/ethnicity subgroups. Our results highlight the importance of improving CKM health clinical management, social support, and policy development.

Our study comprehensively described the prevalence of CKM stages in US adults. One recent study based on NHANES 2015-2020 reported that 26.3% of individuals had at least one CKM condition, whereas 1.5% had three CKM conditions.^4^ However, this study only considered diabetes as a metabolic risk factor. Given the progressive nature of CKM conditions, previous criteria may have concealed other significant subclinical abnormalities. By incorporating a broader definition and its staging approach,^3^ our results confirm the previous findings and reveal an even higher prevalence in the early stage of CKM. Our findings are consistent with those of prior studies.^22, 23^ Distinguished from the previous study, we also found that obesity and metabolic abnormalities were highly prevalent in all CKM stages, which was consistent with the context of the global obesity pandemic.^24^ From a pathogenic perspective, the majority of CKM risk factors stem from the excess and dysfunction of adipose tissue,^25^ which leads to various metabolic dysfunctions, including systematic inflammation and insulin resistance.^26^ Adipose tissue abnormalities also contribute to the development of CVD and its related risk factors, chronic kidney disease, cancer, and diabetes mellitus,^27–29^ leading to the terminal stage of CKM. Improving the recognition and clinical management of excess or dysfunctional adiposity, as well as adopting a proactive approach to control metabolic abnormalities could gain significant health benefits in the prevention of CKM progression.

The new CKM staging system was associated with mortality and life expectancy in US adults in a dose-response relationship. Compared with individuals with no CKM, those in CKM stage 4 result in 15.5 years of life loss at age 50, which could be largely attributable to CV death. Notably, the nonsignificant associations between CKM stage 1 and CV death should be interpreted with caution, as the results were underpowered. Previous studies have examined the associations of CVD risk factors combined or alone with life expectancy.^30–32^ Individuals with obesity ^33^ or impaired kidney function ^34^ had a higher risk of premature death. Compared with diabetes or CKD alone, the comorbidity of CKD and diabetes was also associated with an increased risk of mortality.^35^ Our results are in line with those of prior studies and underscore the significant risk of CKM multimorbidity. We advocate for routine screening, interdisciplinary care, social support, and policy development to enhance CKM health.

There are disparities in prevalence, mortality, and life expectancy among the sex, race/ethnicity, and SDOH subgroups. Compared with males, females exhibited a lower prevalence of high-stage CKM, but a higher risk of mortality and years of life lost. The disparity in prevalence between males and females may be partially attributed to the lower prevalence of CVD diagnostic angiograms in females.^36^ Prior studies have suggested that females may exhibit atypical symptoms and receive less aggressive treatments than males do.^36–39^ This may result in misdiagnosis or delayed diagnosis, as well as health outcome disparities. Other potential factors, including changes in estrogen after menopause,^40, 41^ high prevalence of adverse psychological factors,^42, 43^ disparities in socioeconomic factors,^44^ and low adherence to interventions also contribute to overall health outcomes in females.^45^ Females with kidney failure or CVD were more predisposed to premature death.^46, 47^ We found a higher prevalence of obesity and metabolic disorders among Black and Mexican individuals than among White individuals.^48^ This disparity may be due to a complex interplay of genetic, environmental, socioeconomic, and cultural factors. Black and Mexican individuals are more likely to experience lower socioeconomic status,^49^ food insecurity,^50^ exposure to stressors,^51^ genetic predispositions,^52–54^ and barriers to accessing healthcare,^55^ which consequently lead to higher rates of obesity and metabolic disorders. Prior epidemiological studies reported that Black individuals tend to have a higher prevalence of obesity and hypertension, as well as adverse post-CVD outcomes.^56, 57^ As expected, individuals with high SDOH scores had less impact of higher CKM stages on the risk of death. Limited social resources are intrinsically linked to health care barriers and poor lifestyles, and eventually contribute to adverse health outcomes.^58^ However, these results should be interpreted with caution because of the limited sample size of Black individuals and individuals with high SDOH scores. Equitable and targeted care should be provided to mitigate disparities in sex, race/ethnicity, and SDOH, as well as to improve short-term and long-term clinical outcomes.

Our study had several limitations. First, the NHANES did not collect longitudinal data such as changes in the CKM stages and related conditions. Second, some metabolic indicators were self-reported, which may lead to misclassification and introduce recall bias. Furthermore, the prevalence of subclinical CVD and clinical CVD may be underestimated due to a lack of data on some types of subclinical CVD (NT-proBNP or coronary artery calcification, etc.) and other CVD such as atrial fibrillation and peripheral artery disease. Third, this study consisted of serial cross-sectional survey data when the demographic structure ahead of time may differ from that currently in the US, hence, generalizability may be limited in Asians and other races/ethnicities. Fourth, the number of CV death events in our study was limited, leading to underpowered results in CKM stage 1. Further investigations on CV death in individuals with CKM are warranted. Finally, owing to the observational design, we could not rule out unmeasured confounding factors. For example, the social determinants of health did not include other domains, such as community resources, environmental factors, interpersonal relationships or genetics. These factors may collectively influence the progression of CKM conditions across the life course.

In conclusion, most individuals had CKM stage 1 to 4, especially stage 2 with metabolic disorders and/or CKD. Individuals with a high stage of CKM were associated with an increased risk of mortality and a shorter life expectancy. Our findings suggest the importance of enhancing CKM health screening, interdisciplinary care, social support, and policy development.

## Data Availability

The data described in the article are publicly and freely available without restriction at https://www.cdc.gov/nchs/nhanes/index.htm.

## Abbreviations

CVD: cardiovascular disease
CDC: Centers for Disease Control and Prevention
CI: confidence interval
CKD: chronic kidney disease
CKM: Cardiovascular-Kidney-Metabolic
eGFR: estimated glomerular filtration rate
CV: cardiovascular
HR: hazard ratio
ICD-10: International Classification of Diseases, Tenth Edition
KDIGO: The Kidney Disease: Improving Global Outcomes
MEC: mobile examination center
MetS: metabolic syndrome
NCHS: National Center for Health Statistics
NDI: National Death Index
NHANES: National Health and Nutrition Examination Survey
SDOH: social determinants of health
US: United States
UACR: urinary albumin creatinine ratio
WONDER: Wide-Ranging Online Data for Epidemiologic Research

## Ethics approval and consent to participate

The National Health and Nutrition Examination Survey (NHANES) is a publicly available database approved by the National Center for Health Statistics institutional review board (Protocol #98-12, Protocol #2005-06, Continuation of Protocol #2005-06, Protocol #2011-17, Continuation of Protocol #2011-17, Protocol #2018-01). All participants provided written informed consent when they completed the national survey in the United States. Ethical review and approval were waived for this study since secondary analysis did not require additional institutional review board approval.

## Data Sharing Statement

The data described in the article are publicly and freely available without restriction at https://www.cdc.gov/nchs/nhanes/index.htm.^9^

## CRediT authorship contribution statement

Study design: Jingkuo Li, Lubi Lei

Data collection and analysis: Lubi Lei

Interpretation of data: Jingkuo Li, Lubi Lei, Wei Wang

Drafting of the manuscript: Jingkuo Li, Lubi Lei

Critical revision of the manuscript for important intellectual content: Yanwu Yu, Boxuan Pu, Yue Peng

Study supervision: Lihua Zhang, Yuanlin Guo.

All authors read and approved the final version of the manuscript. Lihua Zhang accepts full responsibility for the finished work, has access to the data, and controls the decision to publish. Our manuscript has not been previously published elsewhere, and is not under consideration by another journal.

