## Supplementary for "Associations of Cardiovascular-Kidney-Metabolic syndrome with premature mortality and life expectancies in US adults: a cohort study"

**eMethods:**

**eMethod 1 Statistical method used for estimating the difference in life expectancy**

We integrated information from three sources to estimate differences in life expectancies between CKM stages (henceforth “exposure groups”):

(1) Age-specific population all-cause mortality rate derived from National Vital Statistics System of National Center for Health Statistics (NCHS) (2019, from 0 to 100 years).
(2) The adjusted HRs of all-cause mortality in each exposure group (stages of CKM) versus the reference, derived from NHANES 1999-2018.
(3) The age-specific population prevalence (categorized in 10-year age groups) of each exposure groups, derived from NHANES 1999-2018.

Population all-cause mortality rates were obtained from the national vital statistics reports (2019, from 0 to 100 years). Age- and sex- specific population mortality rate was used when we assessed the association of CKM with life expectancy in male and female; Age- and race/ethnicity- specific population mortality rate was used when we assessed the association of CKM with life expectancy in White, Black and Mexican participants (Since the population mortality rate for Mexican were not provided, we use the Hispanic population mortality rates instead).

We used Cox regression models to evaluate the associations between stages of CKM and risk of all-cause death, and/or by sex (sex-specific model) or by race/ethnicity (race/ethnicity-specific model). Several covariates were adjusted in these models, including age (20-34, 35-49, 50-64 and ≥ 65 years old), SDOH score and/or sex or race/ethnicity in the corresponding model. The nationally representative prevalence of each CKM stages were calculated to create lifetables. For the population of 80 years onward, the prevalences were assumed to be the same as that in the 70-79-year age group.

The lifetables for each of the 5 exposure groups (CKM stages) were built based on the estimated group- and age-specific death rate (${IR}_{aj}$), which was calculated by weighting (1) the population mortality rate by the corresponding (2) adjusted HR and (3) prevalence. We inferred the age-specific mortality rates appropriate for our reference group ${IR}_{a0}$ as:

$${IR}_{a0}=\frac{{IR}_{a}}{(P_{a0}+\sum_{j=1}^{2} P_{aj}⨯{HR}_{aj})}$$

Where ${IR}_{a}$ is the population mortality rate for age group 𝑎, $P_{aj}$ is the prevalence of exposure group 𝑗, and ${HR}_{aj}$ is the adjusted hazard ratio in comparison of exposure group 𝑗 versus reference group (𝑗 = 0). The age-specific mortality rates in each of the non-reference exposure groups were then inferred in turn by multiplying the age-specific mortality rate for the reference group ${IR}_{a0}$ by the hazard ratios ${HR}_{aj}$.

$${IR}_{aj}={IR}_{a0}⨯{HR}_{aj} (j>0)$$

We built the life table starting at age 50 years and ending at 100 years by single-year age intervals. Survival probability was set of 1 at age 50 years; probability of survival between ages x and x + 1 was calculated based on probability of dying (mortality rate) between ages x and x + 1 assuming that survivor function declines linearly between ages x and x + 1. The life expectancy at any given age was derived by dividing the total person-years that would be lived beyond age x by the number of persons who survived to that age interval.

Finally, the estimated lower survival time (years) due to higher stage of CKM was calculated as the difference in the life expectancy at any given age between the reference group and each of the exposure group.

**eMethod 2 Statistical method used for estimating contributions of specific causes of death to the absolute difference in life expectancy**

We applied Arriaga's decomposition method to estimate the contributions of specific causes of death to the absolute difference in life expectancy between CKM stages. The analytical process consists of two steps:

(1) Decomposition by each single-age group:

$$TE_{x}=\left[ \frac{l_{x}^{1}}{l_{0}}*\left( \frac{L_{x}^{2}}{l_{x}^{2}}-\frac{L_{x}^{1}}{l_{x}^{1}} \right) \right]+\left[ \frac{T_{x+1}^{2}}{l_{0}}*\left( \frac{l_{x}^{1}}{l_{x}^{2}}-\frac{l_{x+1}^{1}}{l_{x+1}^{2}} \right) \right]$$

For the last open-ended age interval, the contribution can be expressed as

$$TE_{x}=\left[ \frac{l_{x}^{1}}{l_{0}}\times\left( \frac{T_{x}^{2}}{l_{x}^{2}}-\frac{T_{x}^{1}}{l_{x}^{1}} \right) \right]$$

TE_x_ is the total contribution of age group x-x+1, l_x_ is the number of individuals alive at age x, l_0_ is the hypothetical cohort size, L_x_ is the number of person-years lived within the single-age interval, T_x_ and T_x+1_ is the total number of person-years lived above age x and x+1, and l_x+1_ is the number of individuals alive at age x+1. "1" represents the reference group, that is participants at stage 0, and "2" represents those at stage 4.

(2) Decomposition by cause of death within each single-age group:

$$TE_{x}^{i}=TE_{x}*\left[ \frac{m_{ⅈ,x}^{2}}{m_{x}^{2}}\frac{-m_{ⅈ,x}^{1}}{-m_{x}^{1}} \right]$$

TE_x^i is the total contribution of age group x-x+1 due to cause i, TE_x_ is the total contribution of age group x-x+1, m_x_ is the all-cause mortality rate of age group x-x+1, m_i,x_ is the specific mortality rate of age group x-x+1 due to cause i. "1" represents the reference group, that is participants at stage 0, and "2" represents those at stage 4.

**eTables: eTable 1 Detailed algorithm of the simplified 10-year CVD risk models**

| Women | **log-Odds** = -3.307728 + 0.7939329 × (age – 55) /10 + 0.0305239 × (TC – HDL-C – 3.5) – 0.1606857 × (HDL-C – 1.3) /0.3 – 0.2394003 × (min(SBP, 110) – 110) /20 + 0.360078 × (max(SBP, 110) – 130) /20 + 0.8667604 × (if diabetes) + 0.5360739 × (if current smoker) + 0.6045917 × (min(eGFR, 60) – 60) / -15 + 0.0433769 × (max(eGFR, 60) – 90) / -15 + 0.3151672 × (if using anti-hypertensive medication) – 0.1477655 × (if using statin) – 0.0663612 × (if using anti-hypertensive medication) × (max(SBP, 110) – 130) /20 + 0.1197879 × (if using statin) × (TC – HDL-C – 3.5) – 0.0819715 × (age – 55) /10 × (TC – HDL-C – 3.5) + 0.0306769 × (age – 55) /10 × (HDL-C – 1.3) /0.3 – 0.0946348 × (age – 55) /10 × (max(SBP, 110) – 130) /20 – 0.27057 × (age – 55) /10 × (if diabetes) – 0.078715 × (age – 55) /10 × (if current smoker) – 0.1637806 × (age – 55) /10 × (min(eGFR, 60) – 60) / -15  **Risk** = exp(log-Odds) / (1 + exp(log-Odds)) |
| --- | --- |
| Men | **log-Odds** = -3.031168 + 0.7688528 × (age – 55) /10 + 0.0736174 × (TC – HDL-C – 3.5) – 0.0954431 × (HDL-C – 1.3) /0.3 – 0.4347345 × (min(SBP, 110) – 110) /20 + 0.3362658 × (max(SBP, 110) – 130) /20 + 0.7692857 × (if diabetes) + 0.4386871 × (if current smoker) + 0.5378979 × (min(eGFR, 60) – 60) / -15 + 0.0164827 × (max(eGFR, 60) – 90) / -15 + 0.288879 × (if using anti-hypertensive medication) – 0.1337349 × (if using statin) – 0.0475924 × (if using anti-hypertensive medication) × (max(SBP, 110) – 130) /20 + 0.150273 × (if using statin) × (TC – HDL-C – 3.5) – 0.0517874 × (age – 55) /10 × (TC – HDL-C – 3.5) + 0.0191169 × (age – 55) /10 × (HDL-C – 1.3) /0.3 – 0.1049477 × (age – 55) /10 × (max(SBP, 110) – 130) /20 – 0.2251948 × (age – 55) /10 × (if diabetes) – 0.0895067 × (age – 55) /10 × (if current smoker) – 0.1543702 × (age – 55) /10 × (min(eGFR, 60) – 60) / -15  **Risk** = exp(log-Odds) / (1 + exp(log-Odds)) |

**Abbreviations**: eGFR: estimated glomerular filtration rate; HDL: high-density lipoprotein cholesterol; SBP: systolic blood pressure; TC: total cholesterol.

**eTable 2 Methods for evaluating each CKM stage**

| CKM stages | Definition | Criterion | Threshold for CKM conditions |
| --- | --- | --- | --- |
| Stage 0: No CKM risk factors | Individuals with normal BMI and waist circumference, normoglycemia, normotension, a normal lipid profile, and no evidence of CKD or subclinical or clinical CVD | All criteria are met | BMI <25 kg/m^2^ (or <23 kg/m^2^ if Asian ancestry) ^a^ |
|  |  |  | Waist circumference <88/102 cm in female/male (or if Asian ancestry <80/90 cm in female/male) ^a^ |
|  |  |  | Fasting blood glucose < 100 mg/dL and HbA1c < 5.7% and without self-reported diagnosis of diabetes, use of insulin, or oral hypoglycemic agents |
|  |  |  | SBP <130 mm Hg and DBP <80 mm Hg without self-reported diagnosis of hypertension or use of antihypertensive medications |
|  |  |  | HDL cholesterol <50/40 mg/dL in female/male and triglycerides < 150 mg/dL |
|  |  |  | Low-risk CKD in KDIGO classification according to eGFR and UACR: UACR < 30 mg/g and eGFR ≥ 60 ml/min/1.73m^2^. |
|  |  |  | Predicted 10-year CVD risk < 20% |
|  |  |  | No history of chronic heart failure, coronary heart disease, heart attack, or stroke |
| Stage 1: Excess or dysfunctional adiposity | Individuals with overweight/obesity, abdominal obesity, or dysfunctional adipose tissue, without the presence of other metabolic risk factors or CKD | Any of the three criteria is met | BMI ≥25 kg/m^2^ (or ≥23 kg/m^2^ if Asian ancestry) |
|  |  |  | Waist circumference ≥88/102 cm in female/male (or if Asian ancestry ≥80/90 cm in female/male) |
|  |  |  | Fasting blood glucose ≥ 100-124 mg/dL or HbA1c ≥ 5.7%-6.4% and without self-reported diagnosis of diabetes, use of insulin, or oral hypoglycemic agents |
|  |  | All criteria are met | SBP <130 mm Hg and DBP <80 mm Hg without self-reported diagnosis of hypertension or use of antihypertensive medications |
|  |  |  | HDL cholesterol <50/40 mg/dL in female/male and triglycerides < 150 mg/dL |
|  |  |  | Low-risk CKD in KDIGO classification according to eGFR and UACR: UACR < 30 mg/g and eGFR ≥ 60 ml/min/1.73m^2^. |
|  |  |  | Predicted 10-year CVD risk < 20% |
|  |  |  | No history of chronic heart failure, coronary heart disease, heart attack, or stroke |
| Stage 2: Metabolic risk factors and CKD | Individuals with metabolic risk factors (hypertriglyceridemia, hypertension, MetS, diabetes), or CKD | Any of the five criteria is met | Triglycerides ≥ 135 mg/dL |
|  |  |  | SBP ≥130 mm Hg or DBP ≥80 mm Hg or self-reported diagnosis of hypertension or use of antihypertensive medications |
|  |  |  | MetS according to waist circumference, HDL cholesterol, triglycerides, blood pressure, and fasting blood glucose ^b^ |
|  |  |  | Fasting blood glucose ≥ 125 mg/dL or HbA1c ≥ 6.5% or self-reported diagnosis of diabetes, use of insulin, or oral hypoglycemic agents |
|  |  |  | Moderate-to-high-risk CKD in KDIGO classification: UACR ≥ 30 mg/g and eGFR ≥ 60 ml/min/1.73m^2^, UACR < 300 mg/g and eGFR ≤ 45-59 ml/min/1.73m^2^, or UACR < 30 mg/g and eGFR ≤ 30-44 ml/min/1.73m^2^. |
|  |  | All criteria are met | No very high-risk CKD in KDIGO classification |
|  |  |  | Predicted 10-year CVD risk < 20% |
|  |  |  | No history of chronic heart failure, coronary heart disease, heart attack, or stroke |
| Stage 3: Subclinical CVD in CKM | Subclinical CVD among individuals with excess/dysfunctional adiposity,  other metabolic risk factors, or CKD | Any of the two criteria is met | Very high-risk CKD in KDIGO classification: UACR ≥ 300 mg/g and eGFR ≤ 45-59 ml/min/1.73m^2^, UACR ≥ 30 mg/g and eGFR ≤ 30-44 ml/min/1.73m^2^, or eGFR ≤ 29 ml/min/1.73m^2^. |
|  |  |  | Predicted 10-year CVD risk ≥ 20% |
|  |  | Any of the eight criteria is met | BMI ≥25 kg/m^2^ (or ≥23 kg/m^2^ if Asian ancestry) |
|  |  |  | Waist circumference ≥88/102 cm in female/male (or if Asian ancestry ≥80/90 cm in female/male) |
|  |  |  | Fasting blood glucose ≥ 100-124 mg/dL or HbA1c ≥ 5.7%-6.4% and without self-reported diagnosis of diabetes, use of insulin, or oral hypoglycemic agents |
|  |  |  | Triglycerides ≥ 135 mg/dL |
|  |  |  | SBP ≥130 mm Hg or DBP ≥80 mm Hg or self-reported diagnosis of hypertension or use of antihypertensive medications |
|  |  |  | MetS according to waist circumference, HDL cholesterol, triglycerides, blood pressure, and fasting blood glucose |
|  |  |  | Fasting blood glucose ≥ 125 mg/dL or HbA1c ≥ 6.5% or self-reported diagnosis of diabetes, use of insulin, or oral hypoglycemic agents |
|  |  |  | Moderate-to-high-risk CKD in KDIGO classification: UACR ≥ 30 mg/g and eGFR ≥ 60 ml/min/1.73m^2^, UACR < 300 mg/g and eGFR ≤ 45-59 ml/min/1.73m^2^, or UACR < 30 mg/g and eGFR ≤ 30-44 ml/min/1.73m^2^. |
|  |  | The criterion is met | No history of chronic heart failure, coronary heart disease, heart attack, or stroke |
| Stage 4: Clinical CVD in CKM | Clinical CVD among individuals with excess/dysfunctional adiposity,  other CKM risk factors, CKD, or very high CKD risk | The criterion is met | History of chronic heart failure, coronary heart disease, heart attack, or stroke |
|  |  | Any of the nine criteria is met | BMI ≥25 kg/m^2^ (or ≥23 kg/m^2^ if Asian ancestry) |
|  |  |  | Waist circumference ≥88/102 cm in female/male (or if Asian ancestry ≥80/90 cm in female/male) |
|  |  |  | Fasting blood glucose ≥ 100-124 mg/dL or HbA1c ≥ 5.7%-6.4% and without self-reported diagnosis of diabetes, use of insulin, or oral hypoglycemic agents |
|  |  |  | Triglycerides ≥ 135 mg/dL |
|  |  |  | SBP ≥130 mm Hg or DBP ≥80 mm Hg or self-reported diagnosis of hypertension or use of antihypertensive medications |
|  |  |  | MetS according to waist circumference, HDL cholesterol, triglycerides, blood pressure, and fasting blood glucose |
|  |  |  | Fasting blood glucose ≥ 125 mg/dL or HbA1c ≥ 6.5% or self-reported diagnosis of diabetes, use of insulin, or oral hypoglycemic agents |
|  |  |  | Moderate-to-high-risk CKD in KDIGO classification: UACR ≥ 30 mg/g and eGFR ≥ 60 ml/min/1.73m^2^, UACR < 300 mg/g and eGFR ≤ 45-59 ml/min/1.73m^2^, or UACR < 30 mg/g and eGFR ≤ 30-44 ml/min/1.73m^2^. |
|  |  |  | Very high-risk CKD in KDIGO classification: UACR ≥ 300 mg/g and eGFR ≤ 45-59 ml/min/1.73m^2^, UACR ≥ 30 mg/g and eGFR ≤ 30-44 ml/min/1.73m^2^, or eGFR ≤ 29 ml/min/1.73m^2^. |

**Abbreviations:** BMI: body mass index; CKD: chronic kidney disease; CKM: cardiovascular-kidney-metabolic; CVD: cardiovascular disease; DBP: diastolic blood pressure; eGFR: estimated glomerular filtration rate; HDL: high-density lipoprotein; KDIGO: The Kidney Disease: Improving Global Outcomes; MetS: metabolic syndrome; SBP: systolic blood pressure; UACR: urinary albumin to creatinine ratio.

^a^ Asian was not listed as a separate race/ethnicity until NAHNES 2011-2012, therefore the uniform threshold for BMI and waist circumference was used in all participants in NHANES 1999-2010.

^b^ MetS is defined by the presence of 3 or more of the following: (1) Waist circumference ≥88/102 cm in female/male (or if Asian ancestry ≥80/90 cm in female/male); (2) HDL cholesterol ≥50/40 mg/dL in female/male; (3) triglycerides ≥150 mg/dL; (4) elevated blood pressure (SBP ≥130 mm Hg or DBP ≥80 mm Hg and/or use of antihypertensive medications); and (5) fasting blood glucose ≥100 mg/dL.

**eTable 3 Detailed definition and threshold of SDOH domains**

| Field | Domain | Categories and Coding | Questions and Definitions |
| --- | --- | --- | --- |
| Economic Stability | Employment | 1: Employed, student, retired  0: Not employed | Participants were asked what type of work they had done in the last week and, if not working, the main reason why. All participants not working were classified as unemployed, except those who responded they were a student or retired were grouped with those reporting employment. |
|  | Poverty/income ratio | 1: PIR ≥ 300%  0: PIR < 300% | Participants were asked their family income and size, and the PIR is the ratio of family income to poverty. The Department of Health and Human Services poverty guidelines were used as the poverty measure to calculate this ratio. These guidelines are issued each year, in the Federal Register, for determining financial eligibility for certain federal programs. The guidelines vary by family size and geographic location. |
|  | Food security | 1: Full food security  0: Marginal, low, or very low | Participants responded to the U.S. Food Security Survey Module questions whether: 1) they were worried if food would run out before there was money to buy more; 2) the food they bought didn’t last and they didn’t have money to get more; 3) they couldn’t afford to eat balanced meals; 4) they had cut the size of meals or skipped meals because there wasn’t enough money for food; 5) if yes to #4, how often meals were cut or skipped; 6) they ate less than they felt they should because there was not enough money to buy food; 7) they were hungry but didn’t eat because they couldn’t afford food; 8) they lost weight because they didn’t have enough money for food; 9) they did not eat for a whole day because there was not enough money for food; 10) if yes to #9, how often they did not eat for the whole day. Levels of food security were classified as follows: full food security, no affirmative responses; marginal food security, 1-2 affirmative responses; low food security, 3-5 affirmative responses; and very low food security, 6-10 affirmative responses. |
| Education Access and Quality | Education level | 1: College or more  0: High school or less | Participants were asked for the highest grade or level of school they completed. The response categories are: less than 9th grade education, 9-11th grade education (includes 12th grade and no diploma), High school graduate, some college or associates degree, and college graduate or higher. |
| Healthcare Access and Quality | Access to healthcare | 1: Routine place to go for healthcare  0: No routine place, or ER/hospital/other | Participants were asked if there is a place they usually go when sick or needing advice about health. If they answered “yes” or “there is more than one place” to this question, they were classified as having a routine place for healthcare. If yes, but the facility is a hospital emergency room, they were classified as not having a routine place for healthcare. |
|  | Health insurance | 1: Private insurance  0: Government or no insurance | Participants were asked whether they are covered by health insurance or some other kind of health care plan. They are subsequently asked if covered by private insurance or several types of government insurance (Medicare, Medi-Gap, Medicaid, military health care, Indian Health Service, state-sponsored health plan, or other government insurance). |
| Neighborhood and Built Environment | Housing instability | 1: Own home  0: Rent or other arrangement | Participants were asked if the home they are living in is owned, being bought, rented, or occupied by some other arrangement. A person was considered to own the home even if they are still paying on a mortgage. |
| Social and Community Context | Marital status | 1: Married or living with a partner  0: Not married nor living with a partner | Participants were asked whether they were married, widowed, divorced, separated, never married, or living with a partner. Those reporting marriage or living with a partner were grouped together. |

**Abbreviations**: ER: emergency room; PIR: poverty/income ratio.

**eTable 4. Sample size and complete characteristics by CKM stages in adults**

| Variables ^a^ | Total  (n=18350) | Stage 0  (n=1873) | Stage 1  (n=3870) | Stage 2  (n=9960) | Stage 3  (n=1103) | Stage 4  (n=1544) |
| --- | --- | --- | --- | --- | --- | --- |
| Age, years (weighted mean (95% CI)) | 46.0 (45.6, 46.5) | 35.0 (34.3, 35.7) | 39.8 (39.1, 40.5) | 47.6 (47.2, 48.1) | 70.7 (69.9, 71.4) | 61.9 (61.1, 62.7) |
| Female, n (weighted %) | 9398 (51.0) | 1203 (65.8) | 2156 (53.4) | 4999 (48.5) | 423 (42.1) | 617 (40.2) |
| Race/ethnicity, n (weighted %) |  |  |  |  |  |  |
| Non-Hispanic White | 7920 (68.9) | 965 (73.9) | 1533 (65.4) | 4180 (68.6) | 471 (68.8) | 771 (73.0) |
| Non-Hispanic Black | 3743 (10.5) | 307 (8.4) | 785 (11.1) | 1994 (10.2) | 284 (14.6) | 373 (12.0) |
| Mexican | 3351 (8.3) | 242 (5.7) | 778 (10.6) | 1958 (8.7) | 187 (5.7) | 186 (4.2) |
| Other Hispanic | 1616 (5.6) | 142 (5.1) | 386 (6.9) | 883 (5.5) | 89 (5.5) | 116 (3.6) |
| Others ^b^ | 1720 (6.7) | 217 (6.9) | 388 (6.1) | 945 (7.0) | 72 (5.4) | 98 (7.2) |
| SDOH |  |  |  |  |  |  |
| SDOH scores (weighted mean (95% CI)) | 5.5 (5.4, 5.6) | 5.6 (5.4, 5.7) | 5.5 (5.4, 5.7) | 5.5 (5.4, 5.6) | 5.4 (5.2, 5.5) | 5.2 (5.1, 5.4) |
| SDOH score<8, n (weighted %) | 16098 (81.7) | 1608 (79.9) | 3341 (79.8) | 8726 (82.0) | 1026 (89.3) | 1397 (85.8) |
| SDOH domains, n (weighted %) |  |  |  |  |  |  |
| Employed | 14192 (81.4) | 1517 (83.6) | 3136 (84.5) | 7558 (80.3) | 929 (86.3) | 1052 (72.8) |
| High PIR | 6572 (48.1) | 776 (52.7) | 1490 (50.3) | 3617 (48.2) | 268 (33.8) | 421 (38.5) |
| Full food security | 13029 (78.2) | 1416 (82.6) | 2709 (78.3) | 7013 (77.4) | 837 (81.2) | 1054 (75.4) |
| College graduate | 9376 (59.6) | 1198 (69.2) | 2244 (65.1) | 4890 (57.4) | 403 (44.9) | 641 (48.3) |
| Routine access to healthcare | 14709 (81.9) | 1360 (76.8) | 2908 (78.4) | 7995 (82.5) | 1025 (93.6) | 1421 (92.7) |
| Private insurance | 9899 (64.3) | 1136 (68.4) | 2189 (66.4) | 5388 (64.5) | 531 (56.6) | 655 (52.6) |
| Own home | 11427 (68.6) | 999 (61.6) | 2244 (65.7) | 6345 (70.0) | 813 (79.4) | 1026 (73.9) |
| Partnered | 11474 (65.9) | 1045 (60.3) | 2397 (65.6) | 6404 (67.2) | 665 (61.1) | 963 (69.4) |
| CKM risk factors |  |  |  |  |  |  |
| HbA1c, % (weighted mean (95% CI)) | 5.57 (5.55, 5.59) | 5.12 (5.10, 5.14) | 5.30 (5.28, 5.31) | 5.66 (5.63, 5.68) | 6.42 (6.32, 6.52) | 6.13 (6.03, 6.22) |
| TC, mg/dL (weighted mean (95% CI)) | 195.08 (194.27, 196.26) | 179.04 (177.17, 180.91) | 187.80 (186.12, 189.49) | 203.97 (202.62, 205.31) | 188.62 (185.25, 191.99) | 184.71 (181.70, 187.72) |
| UACR, mg/g (weighted mean (95% CI)) | 28.70 (25.39, 32.01) | 7.14 (6.84, 7.44) | 6.64 (6.24, 6.64) | 24.96 (21.69, 28.23) | 165.19 (117.27, 213.11) | 102.43 (71.85, 133.02) |
| eGFR, ml/min/1.73m^2^ (weighted mean (95% CI)) | 97.90 (97.37, 98.43) | 106.45 (105.49, 107.41) | 103.51 (102.61, 104.41) | 97.58 (97.01, 98.16) | 69.86 (68.18, 71.54) | 80.19 (78.71, 81.66) |
| History of cancer, n (weighted %) | 1443 (8.5) | 71 (4.7) | 163 (5.2) | 716 (8.2) | 219 (26.4) | 274 (19.7) |
| History of liver disease, n (weighted %) | 729 (3.8) | 32 (1.8) | 83 (1.7) | 444 (4.5) | 50 (5.5) | 120 (7.7) |
| History of lung disease, n (weighted %) | 323 (1.7) | 5 (0.3) | 21 (0.7) | 144 (1.5) | 32 (3.5) | 121 (8.1) |
| CKM conditions |  |  |  |  |  |  |
| CVD, n (weighted %) | 1544 (6.8) | 0 (0.0) | 0 (0.0) | 0 (0.0) | 0 (0.0) | 1544 (100.0) |
| ASCVD | 1384 (6.1) | 0 (0.0) | 0 (0.0) | 0 (0.0) | 0 (0.0) | 1384 (90.2) |
| HF | 484 (2.0) | 0 (0.0) | 0 (0.0) | 0 (0.0) | 0 (0.0) | 484 (29.6) |
| CKD risk, n (weighted %) |  |  |  |  |  |  |
| Moderate | 1905 (8.2) | 0 (0.0) | 0 (0.0) | 1276 (11.2) | 298 (25.4) | 331 (18.7) |
| High | 450 (1.7) | 0 (0.0) | 0 (0.0) | 180 (1.5) | 129 (11.6) | 141 (7.2) |
| Very high | 258 (0.8) | 0 (0.0) | 0 (0.0) | 0 (0.0) | 131 (10.7) | 127 (5.8) |
| Metabolic risk factors, n (weighted %) | 12189 (62.0) | 0 (0.0) | 0 (0.0) | 9679 (96.8) | 1082 (97.6) | 1428 (90.2) |
| Hypertriglyceridemia | 6299 (33.6) | 0 (0.0) | 0 (0.0) | 5072 (53.0) | 536 (49.3) | 691 (46.2) |
| hypertension | 8936 (44.5) | 0 (0.0) | 0 (0.0) | 6617 (66.2) | 1042 (94.8) | 1277 (79.2) |
| MetS | 7338 (37.0) | 0 (0.0) | 0 (0.0) | 5518 (55.4) | 793 (73.3) | 1027 (64.5) |
| Diabetes | 3146 (12.7) | 0 (0.0) | 0 (0.0) | 1815 (15.0) | 677 (58.6) | 654 (37.0) |
| Excess/dysfunctional adiposity, n (weighted %) | 15090 (79.7) | 0 (0.0) | 3870 (100.0) | 8815 (87.4) | 996 (91.3) | 1409 (92.0) |
| Overweight/Obesity | 13138 (69.0) | 0 (0.0) | 3073 (78.4) | 7898 (78.7) | 899 (80.9) | 1268 (82.0) |
| Abdominal obesity | 10589 (55.2) | 0 (0.0) | 2102 (53.5) | 6554 (65.2) | 809 (76.6) | 1124 (73.2) |
| Prediabetes | 7362 (39.5) | 0 (0.0) | 1803 (45.8) | 4677 (46.6) | 298 (29.0) | 584 (40.8) |
| 10-year CVD risk score, % (weighted mean (95% CI)) | 5.2 (5.0, 5.4) | 1.0 (0.9, 1.1) | 1.8 (1.7, 1.9) | 5.1 (5.0, 5.3) | 25.3 (24.8, 25.8) | 14.6 (13.9, 15.3) |

**Abbreviations**: ASCVD: atherosclerotic cardiovascular disease; CI: confidence interval; CKD: chronic kidney disease; CKM: cardiovascular-kidney-metabolic; CVD: cardiovascular disease; eGFR: estimated glomerular filtration rate; HF: heart failure; HbA1c: hemoglobin a1c; MetS: metabolic syndrome; PIR: poverty income ratio; SDOH: social determinants of health; TC: total cholesterol; UACR: urine albumin-creatinine ratio.

^a^ Weighted to be nationally representative.

^b^ Including Asian and multiracial.

**eTable 5. Baseline characteristics of eligible participants who had complete information and who did not**

| Variables | Total  (n=22149) | Completed  (n=18350) | Non-Completed  (n=3799) | P value |
| --- | --- | --- | --- | --- |
| Age, years (mean (SD)) | 47.8 (16.5) | 47.9 (16.5) | 47.5 (16.6) | 0.168 |
| Female, n (%) | 11362 (51.3) | 9398 (51.2) | 1964 (51.7) | 0.600 |
| Race/ethnicity, n (%) |  |  |  | <0.001 |
| Non-Hispanic White | 9351 (42.2) | 7920 (43.2) | 1431 (37.7) |  |
| Non-Hispanic Black | 4634 (20.9) | 3743 (20.4) | 891 (23.5) |  |
| Mexican | 4072 (18.4) | 3351 (18.3) | 721 (19.0) |  |
| Other Hispanic | 1954 (8.8) | 1616 (8.8) | 338 (8.9) |  |
| Others ^a^ | 2138 (9.7) | 1720 (9.4) | 418 (11.0) |  |
| SDOH scores (mean (SD)) | 4.9 (2.1) | 4.9 (2.0) | 4.5 (2.1) | <0.001 |
| SDOH score<8, n (%) | 18922 (88.3) | 16098 (87.7) | 2824 (91.4) | <0.001 |
| SDOH domains, n (%) |  |  |  |  |
| Employed | 16930 (76.4) | 14192 (77.3) | 2738 (72.1) | <0.001 |
| High PIR | 7652 (34.5) | 6572 (35.8) | 1080 (28.4) | <0.001 |
| Full food security | 15292 (70.6) | 13029 (71.0) | 2263 (68.6) | 0.006 |
| College graduate | 11161 (50.4) | 9376 (51.1) | 1785 (47.0) | <0.001 |
| Routine access to healthcare | 17672 (79.8) | 14709 (80.2) | 2963 (78.0) | 0.003 |
| Private insurance | 11741 (53.1) | 9899 (53.9) | 1842 (49.2) | <0.001 |
| Own home | 13412 (61.4) | 11427 (62.3) | 1985 (56.7) | <0.001 |
| Partnered | 13593 (61.9) | 11474 (62.5) | 2119 (58.6) | <0.001 |
| HbA1c, % (mean (SD)) | 5.71 (1.10) | 5.71 (1.09) | 5.72 (1.15) | 0.504 |
| TC, mg/dL (mean (SD)) | 195.30 (42.44) | 195.08 (42.21) | 196.42 (43.61) | 0.085 |
| UACR, mg/g (median (IQR)) | 6.71 (4.42, 12.85) | 6.70 (4.36, 12.73) | 6.81 (4.66, 13.31) | 0.001 |
| eGFR, ml/min/1.73m^2^ (mean (SD)) | 97.02 (21.24) | 96.97 (21.09) | 97.28 (21.98) | 0.433 |
| History of cancer, n (%) | 1714 (7.7) | 1443 (7.9) | 271 (7.1) | 0.134 |
| History of liver disease, n (%) | 908 (4.1) | 729 (4.0) | 179 (4.7) | 0.041 |
| History of lung disease, n (%) | 417 (1.9) | 323 (1.8) | 94 (2.5) | 0.004 |

**Abbreviations**: eGFR: estimated glomerular filtration rate; HbA1c: hemoglobin a1c; IQR: interquartile range; PIR: poverty income ratio; SD: standard deviation; SDOH: social determinants of health; TC: total cholesterol; UACR: urine albumin-creatinine ratio.

^a^ Including Asian and multiracial.

**eTable 6. Relative proportion of different combination of CKM conditions stratified by CKM stage 1-4 in different survey cycles**

| CKM conditions ^b^ | 1999-2002, n (weighted ^a^ %)  n=3028 | 2003-2008, n (weighted %)  n=5350 | 2009-2014, n (weighted %)  n=6270 | 2015-2018, n (weighted %)  n=3702 |
| --- | --- | --- | --- | --- |
| **Stage 1** |  |  |  |  |
| OB only | 351 (72.4) | 582 (58.1) | 763 (53.8) | 371 (41.0) |
| OB + pre-D | 84 (14.1) | 286 (24.9) | 476 (30.0) | 322 (39.9) |
| Pre-D only | 71 (13.5) | 178 (17.0) | 234 (16.2) | 152 (19.1) |
| **Stage 2** |  |  |  |  |
| OB + MR | 637 (37.3) | 1098 (37.2) | 1213 (37.0) | 650 (30.2) |
| OB + pre-D + MR | 513 (29.3) | 964 (34.3) | 1105 (35.8) | 839 (44.0) |
| MR | 260 (17.9) | 278 (10.9) | 284 (8.7) | 114 (5.8) |
| pre-D + MR | 81 (3.9) | 166 (5.5) | 187 (5.2) | 115 (6.2) |
| OB + MR + mh-CKD | 94 (4.5) | 169 (4.4) | 177 (4.1) | 126 (5.7) |
| Others | 139 (7.1) | 255 (7.7) | 323 (9.2) | 173 (8.1) |
| **Stage 3** |  |  |  |  |
| OB + MR + hr-CVD | 61 (35.6) | 94 (31.0) | 109 (31.0) | 77 (37.9) |
| OB + MR + mh-CKD + hr-CVD | 46 (21.0) | 81 (22.0) | 94 (23.5) | 53 (23.6) |
| OB + pre-D + MR + hr-CVD | 22 (12.5) | 37 (10.5) | 33 (9.3) | 26 (14.3) |
| OB + pre-D + MR + mh-CKD + hr-CVD | 12 (6.8) | 30 (9.4) | 31 (8.6) | 13 (5.8) |
| OB + MR + vh-CKD + hr-CVD | 8 (3.5) | 17 (3.3) | 28 (5.7) | 15 (4.3) |
| MR + mh-CKD + hr-CVD | 10 (5.9) | 13 (4.4) | 14 (3.1) | 5 (1.6) |
| pre-D + MR + hr-CVD | 3 (1.7) | 11 (3.8) | 8 (1.8) | 6 (5.0) |
| Others | 23 (13.0) | 46 (15.6) | 54 (17.0) | 23 (7.5) |
| **Stage 4** |  |  |  |  |
| OB + MR + CVD | 53 (27.5) | 81 (20.8) | 103 (21.8) | 79 (25.0) |
| OB + pre-D + MR + CVD | 33 (17.0) | 88 (19.2) | 87 (20.9) | 59 (23.2) |
| OB + MR + mh-CKD + hr-CVD + CVD | 27 (7.5) | 51 (9.1) | 50 (9.0) | 31 (5.2) |
| OB + MR + hr-CVD + CVD | 15 (4.4) | 34 (7.1) | 33 (5.5) | 24 (7.8) |
| OB + MR + mh-CKD + CVD | 17 (6.4) | 31 (5.9) | 35 (5.3) | 31 (7.1) |
| OB + pre-D + MR + mh-CKD + CVD | 9 (2.1) | 23 (5.5) | 24 (5.0) | 21 (5.9) |
| OB + pre-D + CVD | 2 (1.6) | 10 (2.1) | 20 (5.7) | 7 (2.8) |
| MR + CVD | 14 (5.5) | 17 (4.1) | 19 (4.4) | 12 (2.8) |
| Others | 68 (28.0) | 135 (26.2) | 114 (22.4) | 87 (20.2) |

**Abbreviations:** CKM: cardiovascular-kidney-metabolic; CVD: cardiovascular disease; hr-CVD: high-risk cardiovascular disease; mh-CKD: moderate-to-high chronic kidney disease risk; vh-CKD: very high chronic kidney disease risk; MR: metabolic risk factors; OB: obesity, including overweight/obesity or abdominal obesity; Pre-D: prediabetes.

^a^ Weighted to be nationally representative.

^b^ Table only shows the combination with relative prevalence in each stage > 5% among all four survey cycles subgroups.

**eTable 7. Relative proportion of different combination of CKM conditions stratified by CKM stage 1-4 in race/ethnicity subgroups**

| CKM conditions ^b^ | White, n (weighted ^a^ %) | Black, n (weighted %) | Mexican, n (weighted %) |
| --- | --- | --- | --- |
| **Stage 1** |  |  |  |
| OB only | 854 (54.5) | 432 (57.8) | 412 (51.6) |
| OB + pre-D | 409 (28.0) | 249 (30.0) | 261 (34.9) |
| Pre-D only | 270 (17.5) | 104 (12.2) | 105 (13.5) |
| **Stage 2** |  |  |  |
| OB + MR | 1511 (36.9) | 677 (35.8) | 748 (38.1) |
| OB + pre-D + MR | 1459 (35.4) | 631 (31.3) | 712 (37.1) |
| MR | 465 (11.0) | 167 (8.9) | 124 (6.0) |
| OB + MR + mh-CKD | 176 (3.9) | 154 (7.1) | 148 (7.0) |
| OB + pre-D + MR + mh-CKD | 148 (3.2) | 127 (5.7) | 92 (4.6) |
| Pre-D + MR | 238 (5.3) | 123 (5.4) | 68 (3.1) |
| Others | 183 (4.3) | 115 (5.8) | 66 (4.1) |
| **Stage 3** |  |  |  |
| OB + MR + hr-CVD | 154 (35.4) | 72 (24.3) | 63 (35.2) |
| OB + MR + mh-CKD + hr-CVD | 97 (20.2) | 81 (27) | 54 (29.5) |
| OB + MR + vh-CKD + hr-CVD | 15 (2.6) | 29 (11.4) | 13 (5.3) |
| OB + pre-D + MR + hr-CVD | 70 (13.1) | 24 (8.2) | 14 (5.3) |
| OB + pre-D + MR + mh-CKD + hr-CVD | 46 (8.6) | 19 (7.1) | 11 (6.3) |
| Others | 89 (20.1) | 59 (22.0) | 32 (18.4) |
| **Stage 4** |  |  |  |
| OB + MR + CVD | 160 (23.4) | 65 (20.9) | 40 (25.7) |
| OB + pre-D + MR + CVD | 146 (21.3) | 52 (14.1) | 30 (20.5) |
| OB + MR + mh-CKD + hr-CVD + CVD | 72 (7.3) | 43 (9.0) | 27 (11.1) |
| OB + MR + mh-CKD + CVD | 40 (4.9) | 32 (9.1) | 20 (11.7) |
| OB + MR + vh-CKD + hr-CVD + CVD | 22 (2.5) | 31 (7.7) | 7 (3.5) |
| OB + MR + hr-CVD + CVD | 63 (7.4) | 25 (5.0) | 11 (3.2) |
| MR + CVD | 27 (3.9) | 18 (6.0) | 11 (6.4) |
| OB + pre-D + MR + mh-CKD + CVD | 42 (5.3) | 18 (4.6) | 10 (5.3) |
| Others | 199 (24.0) | 89 (23.6) | 30 (12.6) |

**Abbreviations:** CKM: cardiovascular-kidney-metabolic; CVD: cardiovascular disease; hr-CVD: high-risk cardiovascular disease; mh-CKD: moderate-to-high chronic kidney disease risk; MR: metabolic risk factors; OB: obesity, including overweight/obesity or abdominal obesity; Pre-D: prediabetes.

^a^ Weighted to be nationally representative.

^b^ Table only shows the combination with relative prevalence in each stage > 5% among all three race/ethnicity subgroups.

**eTable 8. Associations of CKM stages with all-cause death and cardiovascular death in adults stratified by age subgroups**

|  | All-cause death |  | CV death |  |
| --- | --- | --- | --- | --- |
| CKM stages | Case (%) ^a^ | HR (95% CI) ^b^ | Case (%) | HR (95% CI) |
| 20-34 years |  |  |  |  |
| Stage 0 | 15 (1.3) | Reference | 1 (0.0) | Reference |
| Stage 1 | 12 (0.8) | 0.66 (0.28, 1.56) | 1 (0.0) | 1.33 (0.08, 21.40) |
| Stage 2 | 36 (1.8) | 1.19 (0.59, 2.40) | 12 (0.5) | 13.00 (1.53, 110.00) |
| Stage 3 | 1 (10.8) | 4.66 (0.43, 50.80) | 0 (0.0) | NA |
| Stage 4 | 1 (2.4) | 1.57 (0.13, 18.20) | 0 (0.0) | NA |
| 35-49 years |  |  |  |  |
| Stage 0 | 10 (1.9) | Reference | 0 (0.0) | Reference |
| Stage 1 | 29 (1.9) | 1.25 (0.46, 3.40) | 6 (0.3) | NA |
| Stage 2 | 127 (4.5) | 2.18 (0.96, 4.92) | 32 (1.0) | NA |
| Stage 3 | 9 (55.9) | 46.10 (20.20, 105.00) | 3 (14.4) | NA |
| Stage 4 | 28 (15.2) | 6.33 (2.56, 15.60) | 10 (5.8) | NA |
| 50-64 years |  |  |  |  |
| Stage 0 | 11 (3.9) | Reference | 3 (1.7) | Reference |
| Stage 1 | 56 (4.7) | 1.34 (0.63, 2.86) | 13 (1.1) | 0.66 (0.17, 2.48) |
| Stage 2 | 329 (8.6) | 1.98 (0.91, 4.34) | 73 (1.9) | 0.90 (0.27, 2.97) |
| Stage 3 | 50 (36.1) | 7.55 (2.98, 19.10) | 14 (7.8) | 3.59 (0.94, 13.80) |
| Stage 4 | 156 (28.7) | 5.87 (2.74, 12.60) | 58 (10.8) | 4.66 (1.39, 15.60) |
| 65-79 years |  |  |  |  |
| Stage 0 | 10 (16.9) | Reference | 1 (0.4) | Reference |
| Stage 1 | 42 (14.4) | 0.95 (0.43, 2.12) | 7 (2.3) | 6.40 (0.74, 55.40) |
| Stage 2 | 396 (21.9) | 0.98 (0.49, 1.97) | 109 (5.9) | 10.90 (1.46, 82.20) |
| Stage 3 | 367 (37.5) | 1.90 (0.92, 3.94) | 121 (13.0) | 27.50 (3.62, 210.00) |
| Stage 4 | 369 (41.6) | 2.36 (1.11, 5.03) | 140 (14.9) | 35.90 (4.76, 271.00) |

**Abbreviations:** CI: confidence interval; CKM: cardiovascular-kidney-metabolic; CV: cardiovascular; HR: hazard ratio; NA: not available.

^a^ Weighted to be nationally representative.

^b^ Models were adjusted for sex, race/ethnicity, SDOH score.

**eTable 9. Associations of CKM stages with all-cause death and cardiovascular death in adults stratified by SDOH subgroups**

|  | All-cause death |  | CV death |  |
| --- | --- | --- | --- | --- |
| CKM stages | Case (%) ^a^ | HR (95% CI) ^b^ | Case (%) | HR (95% CI) |
| SDOH score=8 |  |  |  |  |
| Stage 0 | 4 (1.5) | Reference | 2 (0.7) | Reference |
| Stage 1 | 15 (1.8) | 1.28 (0.47, 3.51) | 3 (0.4) | 0.57 (0.09, 3.77) |
| Stage 2 | 73 (4.7) | 2.08 (0.92, 4.70) | 13 (0.7) | 0.52 (0.13, 2.13) |
| Stage 3 | 14 (16.4) | 2.41 (0.81, 7.23) | 5 (6.9) | 1.53 (0.33, 7.01) |
| Stage 4 | 34 (19.7) | 3.72 (1.40, 9.89) | 13 (7.7) | 2.15 (0.47, 9.91) |
| SDOH score<8 |  |  |  |  |
| Stage 0 | 42 (2.3) | Reference | 3 (0.1) | Reference |
| Stage 1 | 124 (2.8) | 1.11 (0.69, 1.78) | 24 (0.5) | 3.36 (0.68, 16.50) |
| Stage 2 | 815 (8.2) | 1.60 (1.13, 2.26) | 213 (2.1) | 7.32 (1.61, 33.30) |
| Stage 3 | 413 (40.2) | 3.65 (2.44, 5.45) | 133 (12.9) | 21.10 (4.34, 103.00) |
| Stage 4 | 520 (34.7) | 4.23 (2.85, 6.26) | 195 (12.6) | 28.20 (5.97, 133.00) |

**Abbreviations:** CI: confidence interval; CKM: cardiovascular-kidney-metabolic; CV: cardiovascular; HR: hazard ratio; SDOH: social determinants of health.

^a^ Weighted to be nationally representative.

^b^ Models were adjusted for sex, age, race/ethnicity.

**eTable 10. Associations of CKM stages with all-cause death and cardiovascular death in adults stratified by race/ethnicity subgroups**

|  | All-cause death |  | CV death |  |
| --- | --- | --- | --- | --- |
| CKM stages | Case (%) ^a^ | HR (95% CI) ^b^ | Case (%) | HR (95% CI) |
| White |  |  |  |  |
| Stage 0 | 26 (2.2) | Reference | 2 (0.2) | Reference |
| Stage 1 | 65 (2.9) | 1.12 (0.63, 1.98) | 13 (0.5) | 2.36 (0.50, 11.20) |
| Stage 2 | 495 (8.7) | 1.73 (1.12, 2.67) | 114 (2) | 4.85 (1.10, 21.30) |
| Stage 3 | 210 (40.5) | 3.50 (2.15, 5.71) | 68 (12.9) | 13.10 (2.86, 60.10) |
| Stage 4 | 328 (35.2) | 4.24 (2.61, 6.89) | 119 (12.5) | 17.70 (3.96, 79.10) |
| Black |  |  |  |  |
| Stage 0 | 12 (3.6) | Reference | 1 (0.1) | Reference |
| Stage 1 | 37 (3.6) | 1.14 (0.61, 2.14) | 7 (0.7) | 6.47 (0.79, 52.80) |
| Stage 2 | 179 (7.3) | 1.22 (0.67, 2.21) | 56 (2.2) | 10.00 (1.34, 74.90) |
| Stage 3 | 108 (36.9) | 3.73 (2.00, 6.94) | 40 (14.3) | 33.50 (4.28, 263.00) |
| Stage 4 | 115 (27.3) | 3.28 (1.85, 5.82) | 48 (11.7) | 35.70 (4.39, 291.00) |
| Mexican |  |  |  |  |
| Stage 0 | 5 (1) | Reference | 1 (0.3) | Reference |
| Stage 1 | 27 (1.7) | 1.73 (0.44, 6.77) | 6 (0.4) | 1.39 (0.15, 12.80) |
| Stage 2 | 142 (3.5) | 1.77 (0.50, 6.21) | 40 (1.1) | 2.24 (0.32, 15.70) |
| Stage 3 | 69 (22.4) | 3.13 (0.81, 12.10) | 19 (4.9) | 2.84 (0.35, 23.40) |
| Stage 4 | 63 (23.9) | 5.41 (1.44, 20.40) | 22 (8.2) | 7.67 (1.03, 57.00) |

**Abbreviations:** CI: confidence interval; CKM: cardiovascular-kidney-metabolic; CV: cardiovascular; HR: hazard ratio.

^a^ Weighted to be nationally representative.

^b^ Models were adjusted for sex, age, SDOH score.

**eTable 11. Associations of CKM stages with all-cause death and cardiovascular death in adults after further adjusted for CKM risk factors**

| Outcomes | Subgroups | HR (95% CI) ^a^ |  |  |  |  |
| --- | --- | --- | --- | --- | --- | --- |
|  |  | Stage 0 | Stage 1 | Stage 2 | Stage 3 | Stage 4 |
| All-cause death |  |  |  |  |  |  |
|  | Overall | Reference | 1.17 (0.75, 1.84) | 1.54 (1.10, 2.17) | 2.53 (1.72, 3.71) | 2.97 (2.02, 4.37) |
|  | Sex subgroups |  |  |  |  |  |
|  | Female | Reference | 1.43 (0.78, 2.64) | 1.80 (1.05, 3.10) | 2.54 (1.44, 4.46) | 3.99 (2.17, 7.34) |
|  | Male | Reference | 0.96 (0.53, 1.75) | 1.29 (0.83, 2.00) | 2.36 (1.42, 3.94) | 2.40 (1.47, 3.90) |
|  | Age subgroups |  |  |  |  |  |
|  | 20-34 years | Reference | 0.62 (0.25, 1.53) | 1.12 (0.50, 2.51) | 10.12 (0.47, 217.34) | 1.24 (0.11, 13.86) |
|  | 35-49 years | Reference | 1.28 (0.48, 3.46) | 1.79 (0.80, 3.99) | 12.81 (4.77, 34.45) | 4.22 (1.70, 10.50) |
|  | 50-64 years | Reference | 1.30 (0.60, 2.80) | 1.70 (0.76, 3.82) | 3.60 (1.35, 9.59) | 3.79 (1.67, 8.60) |
|  | 65-79 years | Reference | 1.08 (0.49, 2.40) | 0.99 (0.49, 2.01) | 1.59 (0.76, 3.30) | 1.86 (0.87, 3.96) |
|  | SDOH score subgroups |  |  |  |  |  |
|  | =8 | Reference | 1.40 (0.52, 3.77) | 2.06 (0.93, 4.55) | 1.81 (0.56, 5.86) | 2.33 (0.82, 6.61) |
|  | <8 | Reference | 1.14 (0.71, 1.83) | 1.45 (1.02, 2.07) | 2.49 (1.65, 3.75) | 2.89 (1.93, 4.32) |
|  | Race/ethnicity subgroups |  |  |  |  |  |
|  | White | Reference | 1.16 (0.66, 2.03) | 1.62 (1.04, 2.52) | 2.57 (1.54, 4.27) | 3.02 (1.84, 4.96) |
|  | Black | Reference | 1.14 (0.61, 2.15) | 1.03 (0.56, 1.91) | 2.13 (1.08, 4.18) | 1.88 (1.02, 3.49) |
|  | Mexican | Reference | 1.71 (0.44, 6.61) | 1.47 (0.43, 5.11) | 1.67 (0.43, 6.56) | 3.01 (0.78, 11.68) |
| CV death |  |  |  |  |  |  |
|  | Overall | Reference | 1.77 (0.57, 5.54) | 2.95 (1.00, 8.72) | 5.90 (1.86, 18.72) | 8.60 (2.85, 26.00) |
|  | Sex subgroups |  |  |  |  |  |
|  | Female | Reference | 1.45 (0.28, 7.52) | 1.77 (0.39, 8.02) | 3.51 (0.71, 17.30) | 7.22 (1.45, 35.94) |
|  | Male | Reference | 2.29 (0.52, 10.18) | 4.34 (1.02, 18.35) | 8.40 (2.01, 35.17) | 11.15 (2.69, 46.23) |
|  | Age subgroups |  |  |  |  |  |
|  | 20-34 years | Reference | 1.17 (0.07, 19.08) | 8.29 (1.00, 68.61) | NA | NA |
|  | 35-49 years | Reference | NA | NA | NA | NA |
|  | 50-64 years | Reference | 0.62 (0.16, 2.36) | 0.68 (0.20, 2.32) | 1.10 (0.26, 4.66) | 2.70 (0.77, 9.45) |
|  | 65-79 years | Reference | 6.39 (0.74, 55.41) | 9.38 (1.24, 71.16) | 17.45 (2.22, 137.01) | 22.51 (2.90, 174.50) |
|  | SDOH score subgroups |  |  |  |  |  |
|  | =8 | Reference | 0.69 (0.11, 4.49) | 0.44 (0.11, 1.70) | 0.91 (0.17, 4.77) | 1.68 (0.42, 6.78) |
|  | <8 | Reference | 3.41 (0.70, 16.70) | 6.25 (1.36, 28.79) | 12.51 (2.49, 62.78) | 18.02 (3.69, 87.99) |
|  | Race/ethnicity subgroups |  |  |  |  |  |
|  | White | Reference | 2.39 (0.50, 11.30) | 4.27 (0.97, 18.89) | 8.54 (1.80, 40.52) | 12.01 (2.60, 55.40) |
|  | Black | Reference | 6.73 (0.82, 55.43) | 8.24 (1.09, 62.13) | 19.55 (2.44, 156.42) | 22.48 (2.74, 184.16) |
|  | Mexican | Reference | 1.43 (0.16, 12.56) | 1.74 (0.25, 11.92) | 1.09 (0.13, 9.13) | 3.07 (0.38, 24.81) |

**Abbreviations:** CI: confidence interval; CKM: cardiovascular-kidney-metabolic; CV: cardiovascular; HR: hazard ratio; NA: not available.

NA: The HR cannot be estimated due to 0 events in the reference or the corresponding stage.

a Models were adjusted for sex, age, race/ethnicity, SDOH score, hemoglobin a1c, total cholesterol, UACR, eGFR and history of cancer, liver disease and lung disease.

**eTable 12. Overall life expectancy according to CKM stages in overall adults and by sex subgroups**

| Life expectancy in single-year age | Overall |  |  |  |  | Female |  |  |  |  | Male |  |  |  |  |
| --- | --- | --- | --- | --- | --- | --- | --- | --- | --- | --- | --- | --- | --- | --- | --- |
|  | Stage 0 | Stage 1 | Stage 2 | Stage 3 | Stage 4 | Stage 0 | Stage 1 | Stage 2 | Stage 3 | Stage 4 | Stage 0 | Stage 1 | Stage 2 | Stage 3 | Stage 4 |
| 50 | 41.4 | 40.0 | 35.8 | 27.6 | 25.9 | 43.4 | 40.2 | 36.5 | 30.6 | 26.5 | 39.0 | 39.7 | 35.3 | 25.1 | 24.9 |
| 51 | 40.5 | 39.1 | 34.9 | 26.9 | 25.2 | 42.5 | 39.3 | 35.6 | 29.8 | 25.8 | 38.1 | 38.8 | 34.4 | 24.4 | 24.2 |
| 52 | 39.6 | 38.2 | 34.1 | 26.1 | 24.4 | 41.6 | 38.4 | 34.7 | 28.9 | 25.0 | 37.3 | 37.9 | 33.6 | 23.7 | 23.5 |
| 53 | 38.7 | 37.3 | 33.2 | 25.4 | 23.7 | 40.7 | 37.4 | 33.8 | 28.1 | 24.2 | 36.4 | 37.1 | 32.8 | 23.0 | 22.8 |
| 54 | 37.8 | 36.5 | 32.4 | 24.6 | 23.0 | 39.7 | 36.5 | 33.0 | 27.3 | 23.5 | 35.6 | 36.2 | 32.0 | 22.4 | 22.2 |
| 55 | 37.0 | 35.6 | 31.6 | 23.9 | 22.3 | 38.8 | 35.7 | 32.1 | 26.5 | 22.7 | 34.7 | 35.4 | 31.2 | 21.7 | 21.6 |
| 56 | 36.1 | 34.7 | 30.7 | 23.2 | 21.7 | 37.9 | 34.8 | 31.2 | 25.7 | 22.0 | 33.9 | 34.6 | 30.4 | 21.1 | 20.9 |
| 57 | 35.2 | 33.9 | 29.9 | 22.5 | 21.0 | 37.0 | 33.9 | 30.4 | 25.0 | 21.3 | 33.1 | 33.7 | 29.6 | 20.5 | 20.4 |
| 58 | 34.4 | 33.0 | 29.1 | 21.9 | 20.4 | 36.1 | 33.0 | 29.6 | 24.2 | 20.6 | 32.3 | 32.9 | 28.9 | 20.0 | 19.8 |
| 59 | 33.5 | 32.2 | 28.4 | 21.2 | 19.8 | 35.2 | 32.1 | 28.7 | 23.5 | 20.0 | 31.5 | 32.1 | 28.1 | 19.4 | 19.2 |
| 60 | 32.7 | 31.4 | 27.6 | 20.6 | 19.2 | 34.3 | 31.3 | 27.9 | 22.7 | 19.3 | 30.7 | 31.3 | 27.4 | 18.9 | 18.7 |
| 61 | 31.8 | 30.5 | 26.8 | 19.9 | 18.5 | 33.4 | 30.4 | 27.1 | 22.0 | 18.6 | 29.9 | 30.5 | 26.6 | 18.3 | 18.1 |
| 62 | 31.0 | 29.7 | 26.0 | 19.3 | 17.9 | 32.5 | 29.5 | 26.2 | 21.2 | 18.0 | 29.1 | 29.7 | 25.9 | 17.7 | 17.5 |
| 63 | 30.1 | 28.9 | 25.2 | 18.6 | 17.3 | 31.7 | 28.7 | 25.4 | 20.5 | 17.3 | 28.3 | 28.9 | 25.1 | 17.1 | 16.9 |
| 64 | 29.3 | 28.0 | 24.4 | 17.9 | 16.7 | 30.8 | 27.8 | 24.6 | 19.8 | 16.6 | 27.5 | 28.1 | 24.3 | 16.5 | 16.3 |
| 65 | 28.4 | 27.2 | 23.6 | 17.3 | 16.1 | 29.9 | 27.0 | 23.8 | 19.0 | 16.0 | 26.7 | 27.3 | 23.6 | 15.9 | 15.8 |
| 66 | 27.6 | 26.4 | 22.9 | 16.7 | 15.5 | 29.0 | 26.1 | 23.0 | 18.3 | 15.4 | 25.9 | 26.5 | 22.9 | 15.4 | 15.2 |
| 67 | 26.8 | 25.6 | 22.1 | 16.1 | 14.9 | 28.1 | 25.3 | 22.2 | 17.6 | 14.7 | 25.1 | 25.7 | 22.1 | 14.8 | 14.7 |
| 68 | 26.0 | 24.8 | 21.4 | 15.4 | 14.3 | 27.3 | 24.4 | 21.4 | 16.9 | 14.1 | 24.3 | 24.9 | 21.4 | 14.3 | 14.2 |
| 69 | 25.2 | 24.0 | 20.6 | 14.9 | 13.7 | 26.4 | 23.6 | 20.6 | 16.2 | 13.5 | 23.6 | 24.1 | 20.7 | 13.8 | 13.7 |
| 70 | 24.4 | 23.2 | 19.9 | 14.3 | 13.2 | 25.6 | 22.8 | 19.9 | 15.6 | 12.9 | 22.8 | 23.4 | 20.0 | 13.3 | 13.2 |
| 71 | 23.5 | 22.3 | 19.1 | 13.6 | 12.6 | 24.7 | 22.0 | 19.0 | 14.8 | 12.3 | 22.0 | 22.5 | 19.2 | 12.7 | 12.6 |
| 72 | 22.7 | 21.5 | 18.3 | 12.9 | 11.9 | 23.8 | 21.1 | 18.2 | 14.1 | 11.6 | 21.2 | 21.7 | 18.4 | 12.0 | 11.9 |
| 73 | 21.8 | 20.7 | 17.5 | 12.3 | 11.3 | 23.0 | 20.3 | 17.5 | 13.4 | 11.0 | 20.4 | 20.9 | 17.7 | 11.4 | 11.3 |
| 74 | 21.0 | 19.9 | 16.8 | 11.6 | 10.7 | 22.1 | 19.5 | 16.7 | 12.7 | 10.4 | 19.6 | 20.1 | 16.9 | 10.8 | 10.7 |
| 75 | 20.2 | 19.1 | 16.0 | 11.0 | 10.1 | 21.3 | 18.6 | 15.9 | 12.1 | 9.8 | 18.8 | 19.3 | 16.2 | 10.2 | 10.1 |
| 76 | 19.4 | 18.3 | 15.3 | 10.4 | 9.5 | 20.4 | 17.8 | 15.1 | 11.4 | 9.2 | 18.0 | 18.5 | 15.4 | 9.6 | 9.5 |
| 77 | 18.6 | 17.5 | 14.6 | 9.8 | 8.9 | 19.6 | 17.1 | 14.4 | 10.7 | 8.6 | 17.2 | 17.7 | 14.7 | 9.1 | 9.0 |
| 78 | 17.8 | 16.8 | 13.8 | 9.2 | 8.4 | 18.8 | 16.3 | 13.7 | 10.1 | 8.1 | 16.5 | 17.0 | 14 | 8.5 | 8.4 |
| 79 | 17.1 | 16.0 | 13.2 | 8.7 | 7.9 | 18.0 | 15.5 | 13.0 | 9.5 | 7.5 | 15.8 | 16.2 | 13.3 | 8.0 | 7.9 |
| 80 | 16.3 | 15.3 | 12.5 | 8.1 | 7.4 | 17.2 | 14.8 | 12.3 | 8.9 | 7.0 | 15.0 | 15.5 | 12.6 | 7.5 | 7.4 |
| 81 | 15.6 | 14.6 | 11.8 | 7.6 | 6.9 | 16.5 | 14.0 | 11.6 | 8.3 | 6.5 | 14.3 | 14.8 | 12.0 | 7.0 | 6.9 |
| 82 | 14.9 | 13.8 | 11.2 | 7.1 | 6.4 | 15.7 | 13.3 | 10.9 | 7.8 | 6.0 | 13.6 | 14.1 | 11.3 | 6.5 | 6.5 |
| 83 | 14.2 | 13.2 | 10.5 | 6.6 | 5.9 | 15.0 | 12.6 | 10.3 | 7.3 | 5.6 | 13.0 | 13.4 | 10.7 | 6.1 | 6.0 |
| 84 | 13.5 | 12.5 | 9.9 | 6.1 | 5.5 | 14.3 | 12 | 9.7 | 6.8 | 5.2 | 12.3 | 12.8 | 10.1 | 5.6 | 5.6 |
| 85 | 12.8 | 11.9 | 9.4 | 5.7 | 5.1 | 13.6 | 11.3 | 9.1 | 6.3 | 4.8 | 11.7 | 12.1 | 9.5 | 5.2 | 5.2 |
| 86 | 12.2 | 11.2 | 8.8 | 5.3 | 4.7 | 12.9 | 10.7 | 8.5 | 5.8 | 4.4 | 11.1 | 11.5 | 9.0 | 4.8 | 4.8 |
| 87 | 11.6 | 10.6 | 8.3 | 4.9 | 4.4 | 12.3 | 10.1 | 8.0 | 5.4 | 4.0 | 10.5 | 10.9 | 8.4 | 4.5 | 4.4 |
| 88 | 11.0 | 10.1 | 7.8 | 4.5 | 4.0 | 11.6 | 9.5 | 7.5 | 5.0 | 3.7 | 9.9 | 10.4 | 7.9 | 4.1 | 4.1 |
| 89 | 10.4 | 9.5 | 7.3 | 4.2 | 3.7 | 11.1 | 9.0 | 7.0 | 4.6 | 3.4 | 9.4 | 9.8 | 7.5 | 3.8 | 3.8 |
| 90 | 9.9 | 9.0 | 6.8 | 3.9 | 3.4 | 10.5 | 8.4 | 6.5 | 4.2 | 3.1 | 8.9 | 9.3 | 7.0 | 3.5 | 3.5 |
| 91 | 9.4 | 8.5 | 6.4 | 3.6 | 3.1 | 10.0 | 8.0 | 6.1 | 3.9 | 2.8 | 8.5 | 8.8 | 6.6 | 3.2 | 3.2 |
| 92 | 8.9 | 8.1 | 6.0 | 3.3 | 2.9 | 9.5 | 7.5 | 5.7 | 3.6 | 2.6 | 8.0 | 8.4 | 6.2 | 3.0 | 3.0 |
| 93 | 8.5 | 7.7 | 5.6 | 3.0 | 2.7 | 9.0 | 7.1 | 5.3 | 3.3 | 2.4 | 7.6 | 8.0 | 5.8 | 2.8 | 2.7 |
| 94 | 8.1 | 7.3 | 5.3 | 2.8 | 2.5 | 8.6 | 6.7 | 5.0 | 3.1 | 2.2 | 7.2 | 7.6 | 5.5 | 2.6 | 2.5 |
| 95 | 7.7 | 6.9 | 5.0 | 2.6 | 2.3 | 8.2 | 6.3 | 4.6 | 2.8 | 2.0 | 6.9 | 7.3 | 5.2 | 2.4 | 2.3 |
| 96 | 7.4 | 6.6 | 4.7 | 2.4 | 2.1 | 7.8 | 6.0 | 4.3 | 2.6 | 1.8 | 6.6 | 7.0 | 4.9 | 2.2 | 2.2 |
| 97 | 7.1 | 6.3 | 4.4 | 2.2 | 1.9 | 7.5 | 5.7 | 4.1 | 2.4 | 1.7 | 6.3 | 6.7 | 4.7 | 2.0 | 2.0 |
| 98 | 6.8 | 6.1 | 4.2 | 2.1 | 1.8 | 7.2 | 5.4 | 3.9 | 2.2 | 1.5 | 6.1 | 6.4 | 4.5 | 1.9 | 1.9 |
| 99 | 6.6 | 5.9 | 4.0 | 1.9 | 1.6 | 7.0 | 5.2 | 3.6 | 2.1 | 1.4 | 5.9 | 6.3 | 4.3 | 1.8 | 1.7 |
| 100 | 6.5 | 5.7 | 3.9 | 1.8 | 1.5 | 6.8 | 5.0 | 3.5 | 1.9 | 1.3 | 5.8 | 6.1 | 4.2 | 1.6 | 1.6 |

**eTable 13. Overall life expectancy according to CKM stages in overall adults and by race/ethnicity subgroups**

| Life expectancy in single-year age | White |  |  |  |  | Black |  |  |  |  | Mexican |  |  |  |  |
| --- | --- | --- | --- | --- | --- | --- | --- | --- | --- | --- | --- | --- | --- | --- | --- |
|  | Stage 0 | Stage 1 | Stage 2 | Stage 3 | Stage 4 | Stage 0 | Stage 1 | Stage 2 | Stage 3 | Stage 4 | Stage 0 | Stage 1 | Stage 2 | Stage 3 | Stage 4 |
| 50 | 41.0 | 39.7 | 35.1 | 27.8 | 25.8 | 39.8 | 38.0 | 37.1 | 23.3 | 24.8 | 44.6 | 38.6 | 38.4 | 32.6 | 27.1 |
| 51 | 40.1 | 38.8 | 34.2 | 27.0 | 25.0 | 39.0 | 37.2 | 36.3 | 22.6 | 24.1 | 43.6 | 37.7 | 37.4 | 31.7 | 26.3 |
| 52 | 39.2 | 37.9 | 33.4 | 26.2 | 24.3 | 38.1 | 36.4 | 35.4 | 22.0 | 23.4 | 42.7 | 36.8 | 36.5 | 30.9 | 25.6 |
| 53 | 38.3 | 37.1 | 32.5 | 25.5 | 23.6 | 37.3 | 35.6 | 34.6 | 21.4 | 22.8 | 41.7 | 35.9 | 35.6 | 30.0 | 24.8 |
| 54 | 37.4 | 36.2 | 31.7 | 24.7 | 22.9 | 36.5 | 34.8 | 33.8 | 20.7 | 22.1 | 40.8 | 35.0 | 34.8 | 29.2 | 24.0 |
| 55 | 36.5 | 35.3 | 30.9 | 24.0 | 22.2 | 35.7 | 34.0 | 33.1 | 20.2 | 21.5 | 39.9 | 34.1 | 33.9 | 28.4 | 23.3 |
| 56 | 35.7 | 34.5 | 30.1 | 23.3 | 21.6 | 34.9 | 33.2 | 32.3 | 19.6 | 20.9 | 39.0 | 33.2 | 33.0 | 27.6 | 22.5 |
| 57 | 34.8 | 33.6 | 29.3 | 22.7 | 20.9 | 34.1 | 32.4 | 31.5 | 19.1 | 20.4 | 38.1 | 32.3 | 32.1 | 26.8 | 21.8 |
| 58 | 33.9 | 32.8 | 28.5 | 22.0 | 20.3 | 33.3 | 31.7 | 30.8 | 18.5 | 19.8 | 37.2 | 31.5 | 31.3 | 26.0 | 21.1 |
| 59 | 33.1 | 31.9 | 27.7 | 21.3 | 19.7 | 32.6 | 30.9 | 30.1 | 18.0 | 19.3 | 36.3 | 30.6 | 30.4 | 25.2 | 20.4 |
| 60 | 32.3 | 31.1 | 26.9 | 20.7 | 19.1 | 31.8 | 30.2 | 29.4 | 17.6 | 18.8 | 35.4 | 29.8 | 29.6 | 24.4 | 19.7 |
| 61 | 31.4 | 30.3 | 26.1 | 20.0 | 18.4 | 31.1 | 29.5 | 28.6 | 17.0 | 18.3 | 34.5 | 28.9 | 28.7 | 23.7 | 19.0 |
| 62 | 30.5 | 29.4 | 25.3 | 19.3 | 17.8 | 30.3 | 28.7 | 27.9 | 16.5 | 17.7 | 33.6 | 28.1 | 27.9 | 22.9 | 18.4 |
| 63 | 29.7 | 28.6 | 24.6 | 18.7 | 17.2 | 29.5 | 28.0 | 27.2 | 16.0 | 17.2 | 32.7 | 27.3 | 27.1 | 22.1 | 17.7 |
| 64 | 28.8 | 27.7 | 23.8 | 18.0 | 16.5 | 28.8 | 27.3 | 26.5 | 15.6 | 16.7 | 31.8 | 26.4 | 26.2 | 21.4 | 17.1 |
| 65 | 28.0 | 26.9 | 23.0 | 17.4 | 15.9 | 28.1 | 26.6 | 25.8 | 15.1 | 16.2 | 30.9 | 25.6 | 25.4 | 20.6 | 16.4 |
| 66 | 27.2 | 26.1 | 22.2 | 16.7 | 15.3 | 27.4 | 25.9 | 25.1 | 14.7 | 15.7 | 30.1 | 24.8 | 24.6 | 19.9 | 15.8 |
| 67 | 26.3 | 25.3 | 21.5 | 16.1 | 14.7 | 26.6 | 25.2 | 24.4 | 14.2 | 15.3 | 29.2 | 24.0 | 23.8 | 19.2 | 15.2 |
| 68 | 25.5 | 24.5 | 20.7 | 15.5 | 14.1 | 25.9 | 24.5 | 23.8 | 13.8 | 14.8 | 28.3 | 23.2 | 23.0 | 18.5 | 14.5 |
| 69 | 24.7 | 23.7 | 20.0 | 14.8 | 13.6 | 25.3 | 23.8 | 23.1 | 13.5 | 14.4 | 27.5 | 22.4 | 22.2 | 17.8 | 13.9 |
| 70 | 23.9 | 22.9 | 19.3 | 14.3 | 13.0 | 24.6 | 23.2 | 22.5 | 13.1 | 14.0 | 26.6 | 21.6 | 21.4 | 17.1 | 13.4 |
| 71 | 23.0 | 22.0 | 18.5 | 13.6 | 12.4 | 23.8 | 22.4 | 21.7 | 12.5 | 13.4 | 25.8 | 20.8 | 20.6 | 16.3 | 12.7 |
| 72 | 22.2 | 21.2 | 17.7 | 12.9 | 11.7 | 23.0 | 21.7 | 20.9 | 12.0 | 12.8 | 24.9 | 20.0 | 19.8 | 15.6 | 12.1 |
| 73 | 21.4 | 20.4 | 16.9 | 12.2 | 11.1 | 22.2 | 20.9 | 20.2 | 11.4 | 12.2 | 24.1 | 19.2 | 19.0 | 14.9 | 11.5 |
| 74 | 20.6 | 19.6 | 16.2 | 11.6 | 10.5 | 21.5 | 20.2 | 19.5 | 10.9 | 11.7 | 23.2 | 18.4 | 18.2 | 14.2 | 10.9 |
| 75 | 19.7 | 18.8 | 15.4 | 11.0 | 9.9 | 20.7 | 19.4 | 18.7 | 10.3 | 11.1 | 22.4 | 17.6 | 17.4 | 13.5 | 10.3 |
| 76 | 18.9 | 18.0 | 14.7 | 10.4 | 9.3 | 20.0 | 18.7 | 18.0 | 9.8 | 10.6 | 21.5 | 16.8 | 16.7 | 12.8 | 9.7 |
| 77 | 18.2 | 17.2 | 14.0 | 9.8 | 8.8 | 19.3 | 18.0 | 17.3 | 9.3 | 10.0 | 20.7 | 16.1 | 15.9 | 12.1 | 9.1 |
| 78 | 17.4 | 16.5 | 13.3 | 9.2 | 8.2 | 18.5 | 17.3 | 16.6 | 8.8 | 9.5 | 19.9 | 15.3 | 15.2 | 11.5 | 8.5 |
| 79 | 16.6 | 15.7 | 12.6 | 8.6 | 7.7 | 17.8 | 16.6 | 16.0 | 8.3 | 9.0 | 19.1 | 14.6 | 14.4 | 10.8 | 8.0 |
| 80 | 15.9 | 15.0 | 11.9 | 8.1 | 7.2 | 17.2 | 15.9 | 15.3 | 7.8 | 8.5 | 18.3 | 13.9 | 13.7 | 10.2 | 7.5 |
| 81 | 15.2 | 14.3 | 11.3 | 7.6 | 6.7 | 16.5 | 15.3 | 14.6 | 7.4 | 8.0 | 17.6 | 13.2 | 13.0 | 9.6 | 7.0 |
| 82 | 14.4 | 13.6 | 10.7 | 7.1 | 6.3 | 15.8 | 14.6 | 14.0 | 7.0 | 7.6 | 16.8 | 12.5 | 12.3 | 9.0 | 6.5 |
| 83 | 13.7 | 12.9 | 10.0 | 6.6 | 5.8 | 15.2 | 14.0 | 13.4 | 6.6 | 7.2 | 16.1 | 11.8 | 11.7 | 8.4 | 6.0 |
| 84 | 13.1 | 12.2 | 9.5 | 6.1 | 5.4 | 14.6 | 13.4 | 12.8 | 6.2 | 6.7 | 15.3 | 11.2 | 11.0 | 7.9 | 5.6 |
| 85 | 12.4 | 11.6 | 8.9 | 5.7 | 5.0 | 14.0 | 12.8 | 12.2 | 5.8 | 6.3 | 14.6 | 10.5 | 10.4 | 7.4 | 5.2 |
| 86 | 11.8 | 11.0 | 8.3 | 5.3 | 4.6 | 13.4 | 12.3 | 11.7 | 5.4 | 6.0 | 14.0 | 9.9 | 9.8 | 6.9 | 4.8 |
| 87 | 11.2 | 10.4 | 7.8 | 4.9 | 4.2 | 12.8 | 11.7 | 11.2 | 5.1 | 5.6 | 13.3 | 9.4 | 9.2 | 6.4 | 4.4 |
| 88 | 10.6 | 9.8 | 7.3 | 4.5 | 3.9 | 12.3 | 11.2 | 10.6 | 4.8 | 5.2 | 12.7 | 8.8 | 8.7 | 6.0 | 4.0 |
| 89 | 10.0 | 9.3 | 6.9 | 4.1 | 3.6 | 11.8 | 10.7 | 10.2 | 4.5 | 4.9 | 12.1 | 8.3 | 8.2 | 5.5 | 3.7 |
| 90 | 9.5 | 8.8 | 6.4 | 3.8 | 3.3 | 11.3 | 10.2 | 9.7 | 4.2 | 4.6 | 11.5 | 7.8 | 7.7 | 5.1 | 3.4 |
| 91 | 9.0 | 8.3 | 6.0 | 3.5 | 3.0 | 10.8 | 9.8 | 9.3 | 3.9 | 4.3 | 11.0 | 7.3 | 7.2 | 4.8 | 3.1 |
| 92 | 8.6 | 7.8 | 5.6 | 3.3 | 2.8 | 10.4 | 9.4 | 8.8 | 3.7 | 4.1 | 10.4 | 6.9 | 6.8 | 4.4 | 2.9 |
| 93 | 8.1 | 7.4 | 5.3 | 3.0 | 2.6 | 10.0 | 9.0 | 8.5 | 3.4 | 3.8 | 10.0 | 6.5 | 6.4 | 4.1 | 2.7 |
| 94 | 7.7 | 7.0 | 4.9 | 2.8 | 2.4 | 9.6 | 8.6 | 8.1 | 3.2 | 3.6 | 9.5 | 6.1 | 6.0 | 3.8 | 2.4 |
| 95 | 7.4 | 6.7 | 4.6 | 2.6 | 2.2 | 9.2 | 8.2 | 7.8 | 3.0 | 3.3 | 9.1 | 5.7 | 5.6 | 3.5 | 2.2 |
| 96 | 7.0 | 6.4 | 4.4 | 2.4 | 2.0 | 8.9 | 7.9 | 7.4 | 2.8 | 3.1 | 8.7 | 5.4 | 5.3 | 3.3 | 2.1 |
| 97 | 6.7 | 6.1 | 4.1 | 2.2 | 1.9 | 8.6 | 7.6 | 7.2 | 2.6 | 2.9 | 8.4 | 5.1 | 5.0 | 3.0 | 1.9 |
| 98 | 6.5 | 5.8 | 3.9 | 2.0 | 1.7 | 8.4 | 7.4 | 6.9 | 2.5 | 2.8 | 8.1 | 4.9 | 4.8 | 2.8 | 1.7 |
| 99 | 6.3 | 5.6 | 3.7 | 1.9 | 1.6 | 8.1 | 7.2 | 6.7 | 2.3 | 2.6 | 7.9 | 4.6 | 4.5 | 2.6 | 1.6 |
| 100 | 6.1 | 5.5 | 3.5 | 1.7 | 1.4 | 8.0 | 7.0 | 6.5 | 2.1 | 2.4 | 7.7 | 4.4 | 4.3 | 2.5 | 1.4 |

**eFigure:**

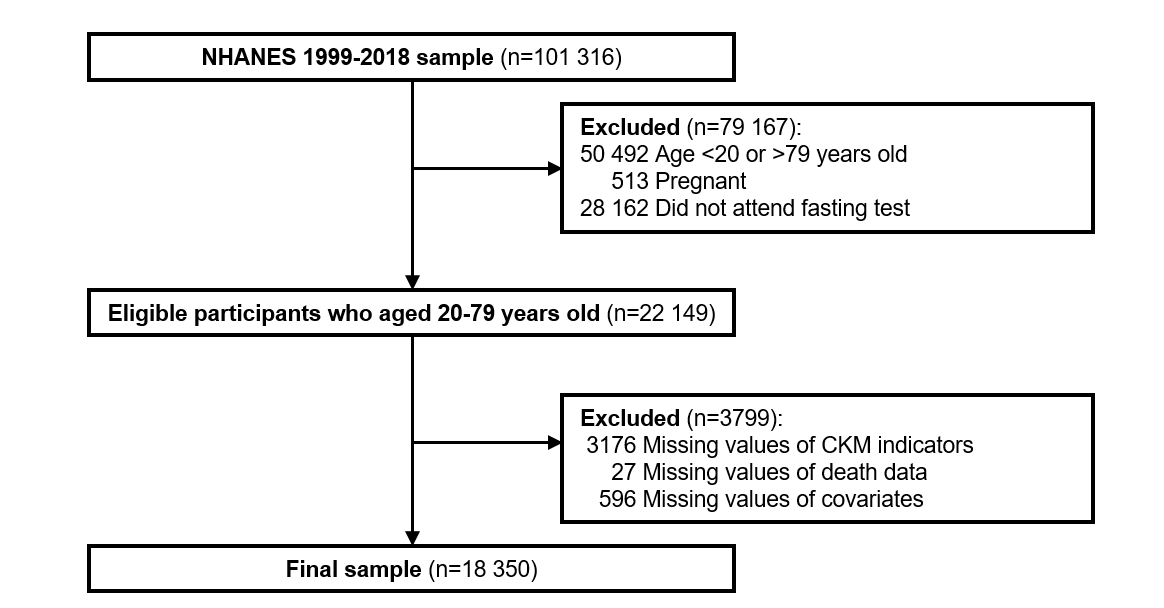

**eFigure 1**. Flow chart of the study sample.

**Abbreviations:** CKM: cardiovascular-kidney-metabolic.

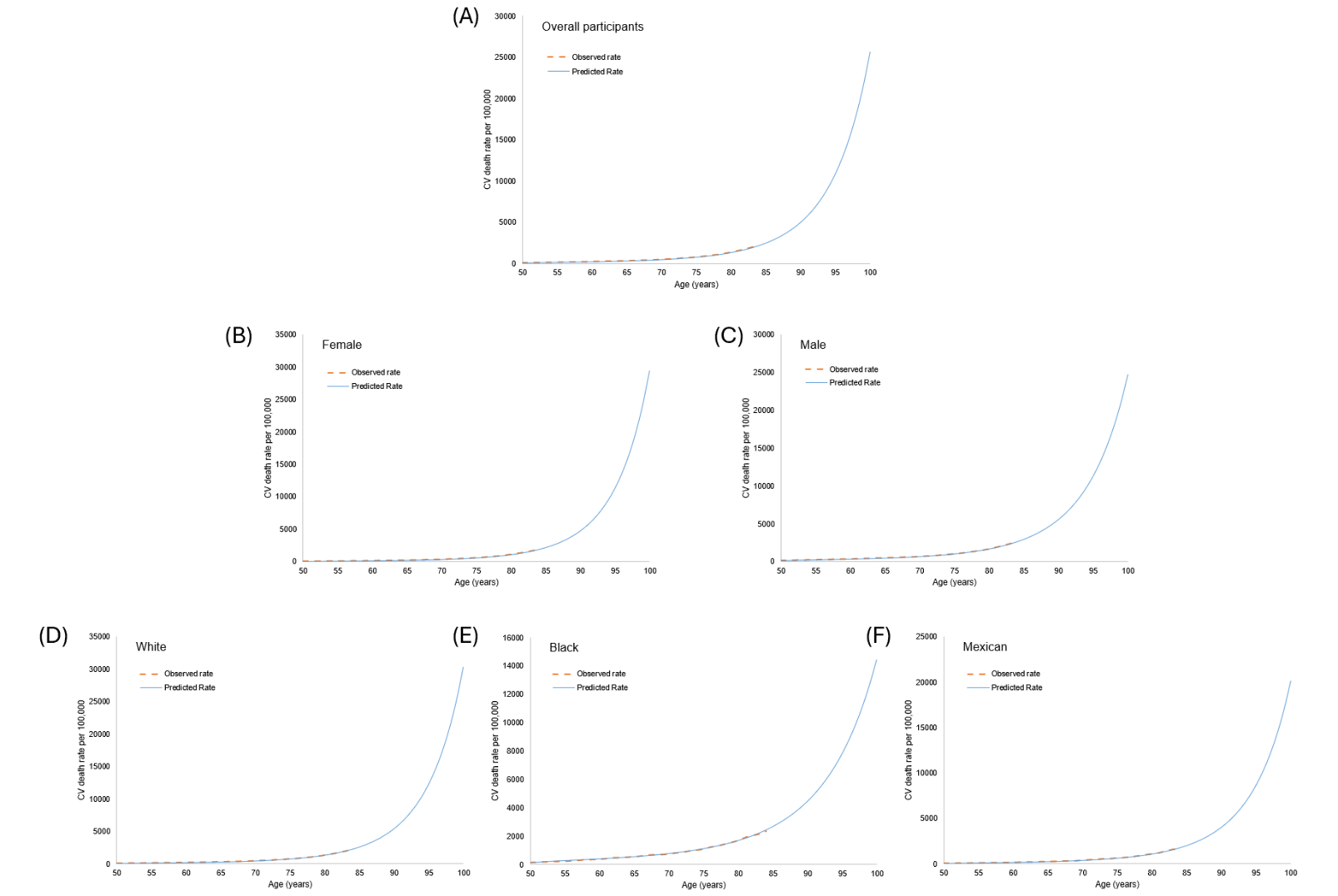

**eFigure 2**. Observed and predicted rate of CV death rates of 2019. (A) Overall; (B) Female; (C) Male; (D) White; (E) Black; (F) Mexican

Abbreviations: CV: cardiovascular.

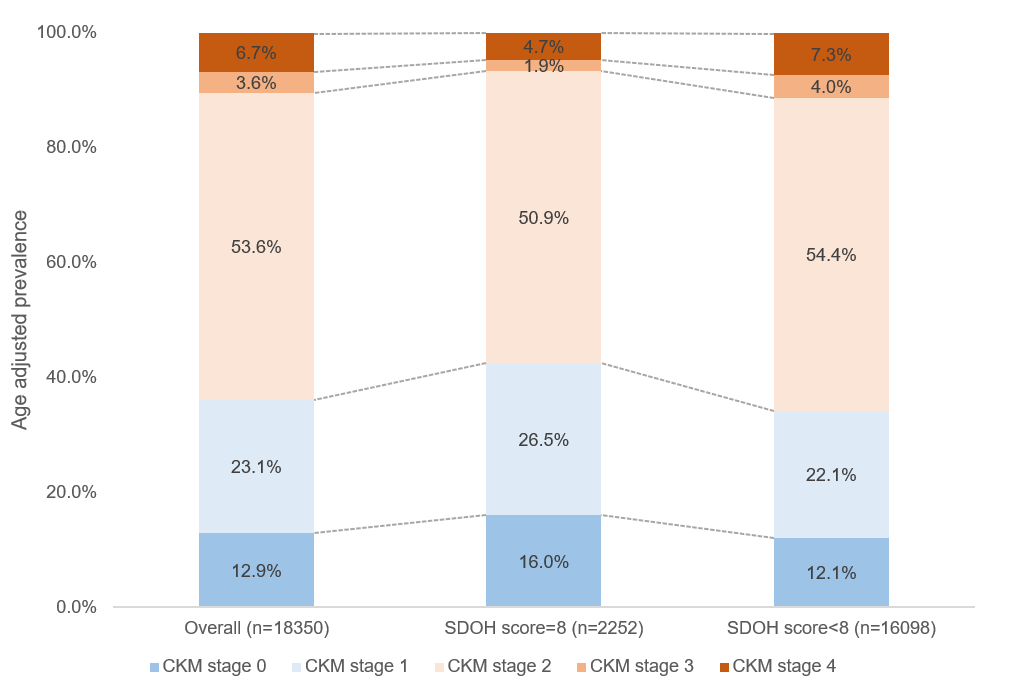

**eFigure 3**. Age adjusted prevalence of CKM stages in overall adults and by SDOH subgroups.

Abbreviations: CKM: cardiovascular-kidney-metabolic; SDOH: social determinants of health.

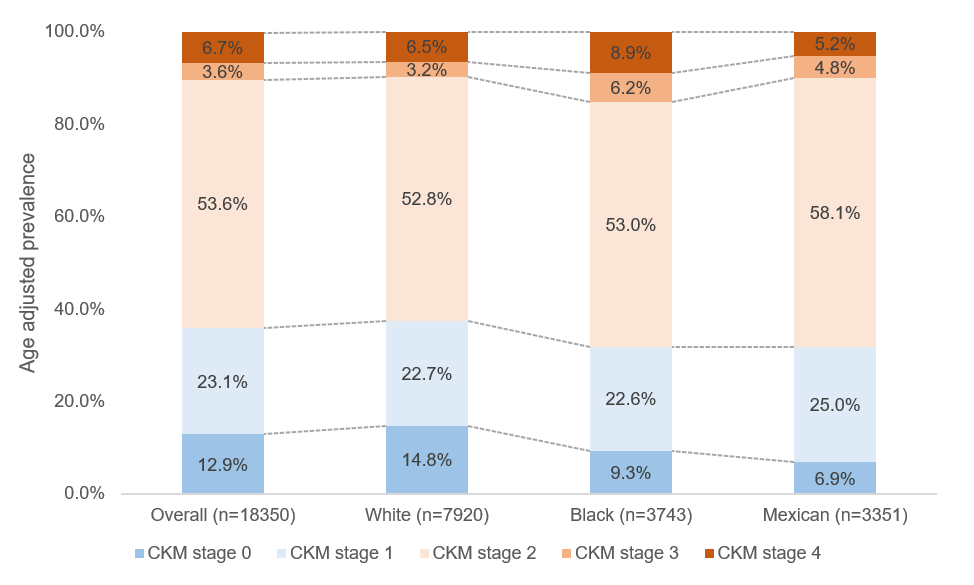

**eFigure 4**. Age adjusted prevalence of CKM stages in overall adults and by race/ethnicity subgroups.

Abbreviations: CKM: cardiovascular-kidney-metabolic.

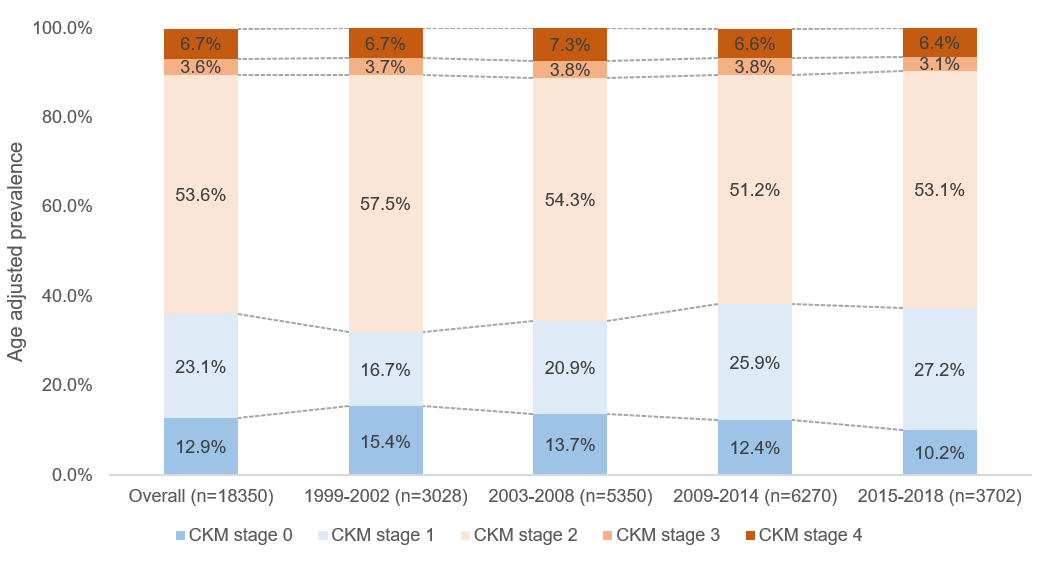

**eFigure 5**. Age adjusted prevalence of CKM stages in overall adults and by survey cycle subgroups.

Abbreviations: CKM: cardiovascular-kidney-metabolic.

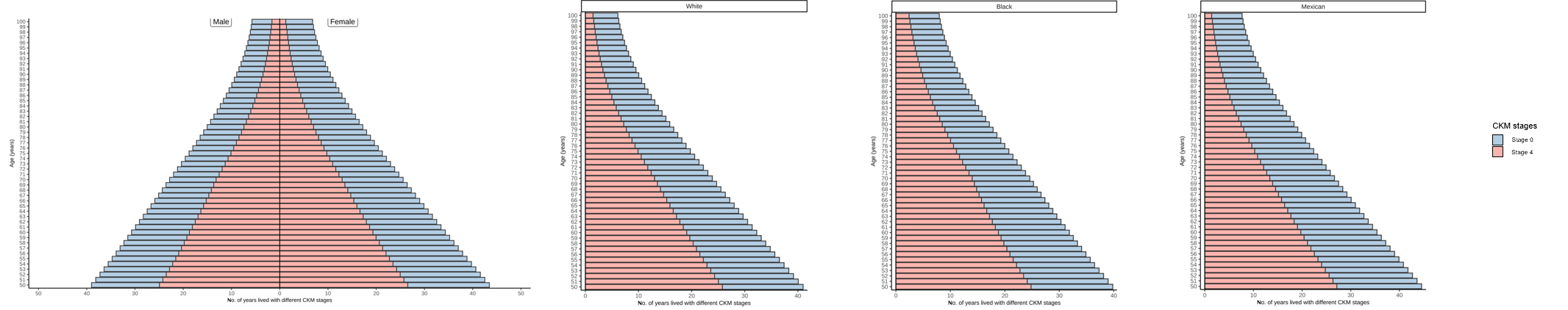

**eFigure 6.** Overall life expectancy between CKM stage 0 and 4 by sex and race/ethnicity subgroups

**Abbreviations:** CKM: cardiovascular-kidney-metabolic.

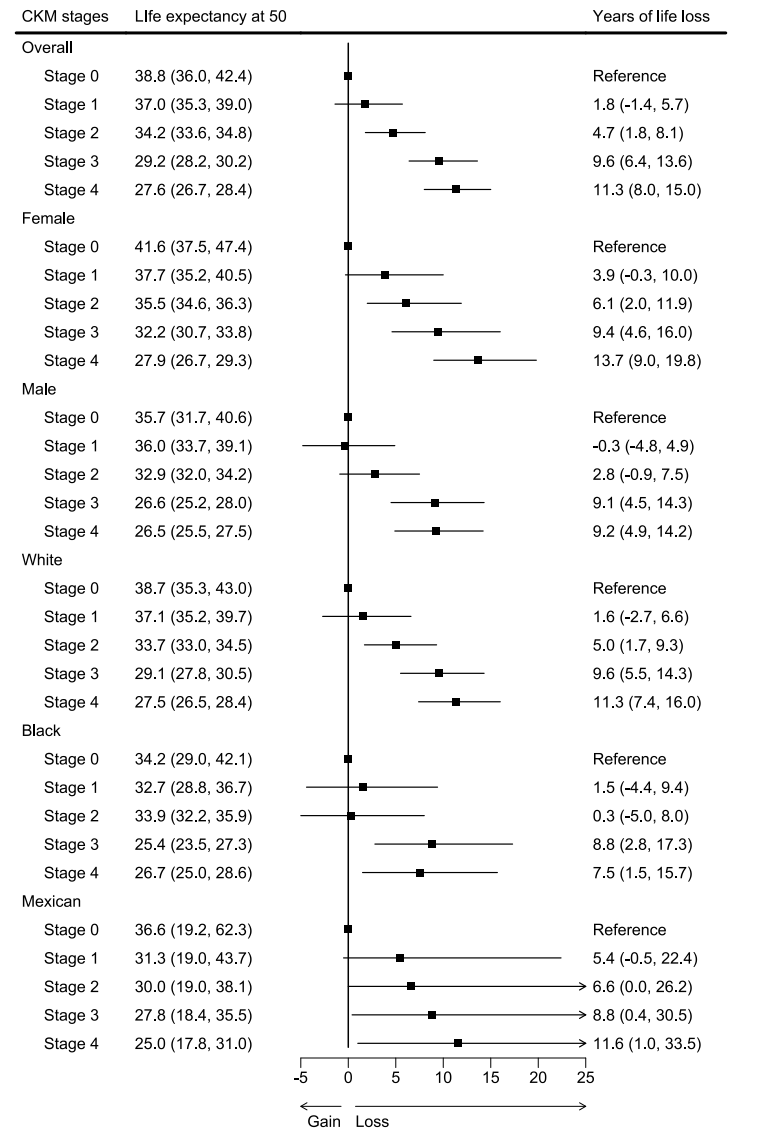

**eFigure 7.** Life expectancy and years of life loss at age 50 according to CKM stages in overall adults and by sex and race/ethnicity subgroups after further adjusting for CKM risk factors

**Abbreviations:** CKM: cardiovascular-kidney-metabolic

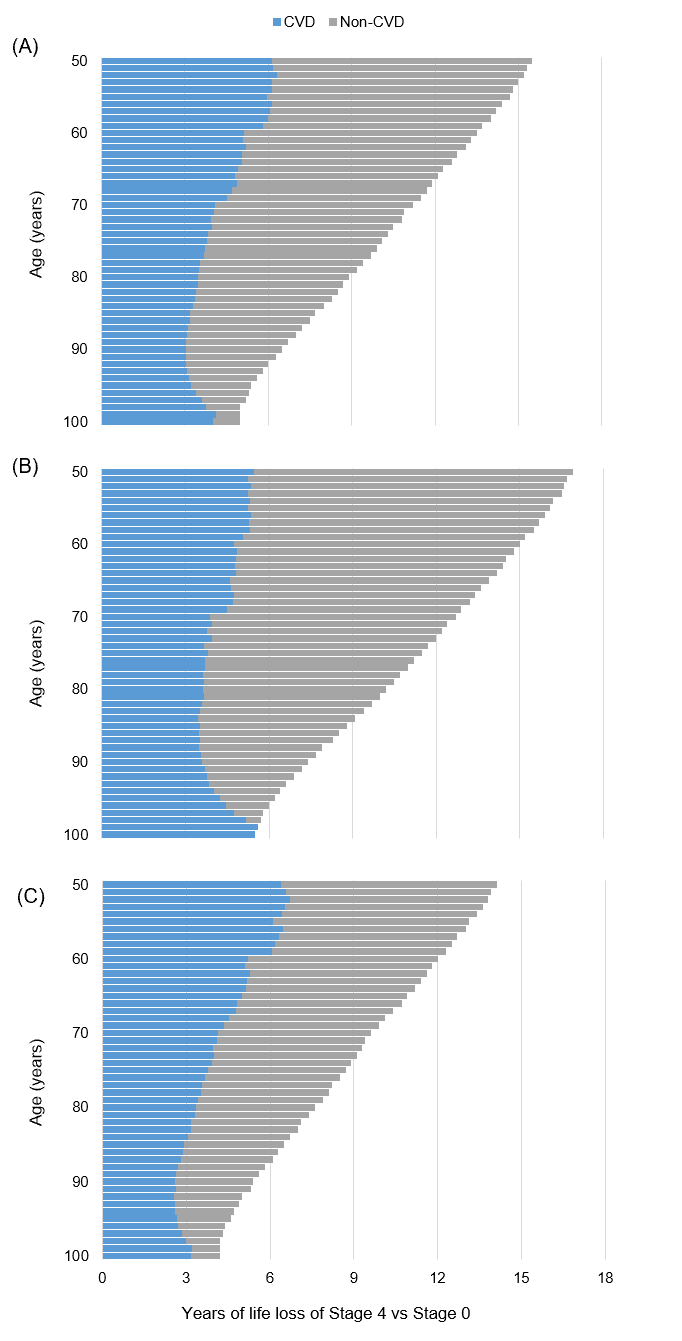

**eFigure 8.** Estimated years of life loss from stage 4 versus stage 0 attributable to increased death from CVD and non-CVD in overall adults and by sex subgroups. (A) Overall participants; (B) Female; (C) Male

**Abbreviations:** CVD: cardiovascular disease.

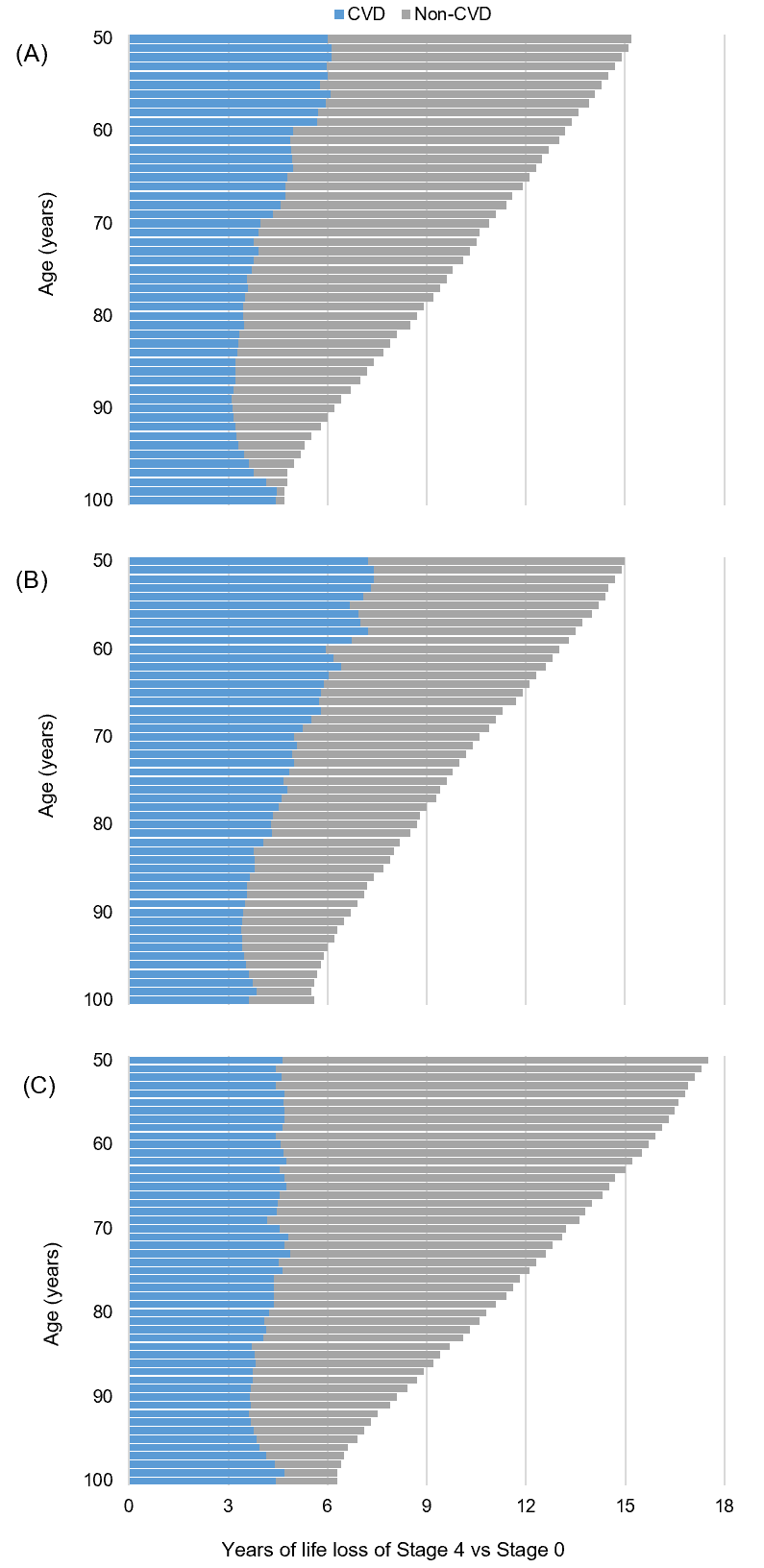

**eFigure 9**. Estimated years of life loss from stage 4 versus stage 0 attributable to increased death from cardiovascular disease and other causes by race/ethnicity subgroups. (A) White; (B) Black; (C) Mexican

Abbreviations: CVD: cardiovascular disease.
